# Estimating the contribution of coding mutations to autism

**DOI:** 10.64898/2026.08.25.26361328

**Authors:** Ajay Nadig, Jack Fu, F. Kyle Satterstrom, Chiara Auwerx, Zhancheng Zhang, Rebecca Torene, Wenhan Lu, Konrad J. Karczewski, The Autism Sequencing Consortium, GeneDx, Joseph D. Buxbaum, Paul Kruszka, Michael Talkowski, Elise B. Robinson, Luke J. O’Connor

## Abstract

*De novo* mutations in protein-coding regions are strongly associated with autism, and family-based sequencing studies have identified numerous genes that harbor excess mutations in probands. However, the aggregate contribution of this class of variation to autism remains unclear. Here, we model the distribution of *de novo* autosomal coding variant effect sizes in 38,680 autism trios to estimate fundamental features of *de novo* genetic architecture. We find that damaging *de novo* single-nucleotide variants and frameshift indels explain 3.4% (95% CI: 2.1% - 4.7%) of autism variance on the observed scale. Approximately 7.0% (95% CI: 5.6% - 8.4%) of cases carry a large-effect mutation (rate ratio > 5), and most such mutations are incompletely penetrant. Although hundreds of genes make some nonzero contribution, 50% of mutational variance on the autosomes is explained by just 15 genes. *De novo* enrichments vary across cohorts with different ascertainment strategies; making projections for future trio studies, we show that many large-effect genes remain to be found.

## Introduction

The autism spectrum is a family of behavioral phenotypes that are developmental in origin and include social deficits and repetitive/restrictive behaviors and interests (Lord *et al*., 2020). Autism is highly heritable (Bai *et al*., 2019), motivating genetic association studies of varied design (Dias and Walsh, 2020). While common variants account for the majority of autism heritability (Gaugler *et al*., 2014; Havdahl *et al*., 2021), and inherited rare variants play a role in autism etiology, *de novo* variants have to date provided the most compelling biological insight into the origins of autism (Fu *et al*., 2022; Zhou *et al*., 2022). This is explained by the fact that autism is associated with large decreases in fecundity (Power *et al*., 2013); this prevents new, large-effect mutations from rising to high allele frequency in the population. *De novo* mutations can be identified in family-based studies where genetic data from both parents are accessible, and these studies have yielded numerous high confidence “autism genes” (Fu *et al*., 2022), illuminated connections with attention-deficit hyperactivity disorder (Satterstrom *et al*., 2019) and the broader family of developmental disorders (Satterstrom *et al*., 2020), highlighted neuronal communication and genome regulation as key biological pathways (De Rubeis *et al*., 2014; Sanders, 2015), and led to genetic diagnoses for thousands of individuals (J. R. Wright *et al*., 2024).

These findings raise questions about what remains to be discovered. What proportion of phenotypic variance is accounted for by *de novo* variation in aggregate? What proportion of individuals with autism carry a likely-causal variant? What is the penetrance spectrum of autism-associated *de novo* mutations?

To answer these questions, we would ideally estimate genetic features of autism in a dataset of infinite sample size. While this is impossible, we can approximate such quantities by fitting a probabilistic model to the observed pattern of effect sizes. Such approaches are ubiquitous in association studies of inherited variation, ranging from estimators of heritability (Yang *et al*., 2010; Bulik-Sullivan *et al*., 2015; Weiner *et al*., 2023) to methods that estimate the full effect size distribution across single-nucleotide polymorphisms (SNPs) (Zhang *et al*., 2018; O’Connor, 2021). An analogous approach for *de novo* variation has the potential to give similar insight into the biology of autism, and, more generally, to provide a deeper quantitative understanding of the genetic architecture of new mutations.

Previous work has characterized aspects of *de novo* genetic architecture. Sanders et al (Sanders *et al*., 2012) estimated that there are approximately 1000 autism genes using methods from quantitative ecology; this estimate was corroborated by He et al (He *et al*., 2013), who fit a univariate gamma model to the distribution of burden effect sizes. Through simulations, Neale et al (Neale *et al*., 2012) clarified that trio sequencing data are inconsistent with a model where most individuals with autism carry highly penetrant *de novo* mutations and consistent with a liability-scale heritability of 1-2.4%. In agreement with this observation, Gaugler et al (Gaugler *et al*., 2014) estimated that *de novo* variants explain 2.6% of autism phenotypic variance on the liability scale (much less than the common variant contribution), assuming that all autism-causing *de novo* variants are fully penetrant. Although these estimates provide key intuition about the nature of *de novo* contributions to autism, they are limited in that they do not precisely characterize the penetrance spectrum of autism, which crucially impacts how mutations in these genes are distributed in the population. In the decade since these estimates were published, sample sizes have increased by an order of magnitude, and new statistical approaches have emerged for flexibly characterizing effect size distributions (Stephens, 2016). These advances present an opportunity to substantially deepen our understanding of the *de novo* genetic architecture of autism.

To characterize the *de novo* genetic architecture of autism, we estimated the distribution of *de novo* burden associations in a dataset of 38,680 autism proband trios. From this inferred distribution, we compute several features of the genetic architecture of autism, investigate how it varies across the autism spectrum, and discuss its clinical implications.

## Results

### Estimating the *de novo* effect size distribution

We analyze exome sequencing data from 38,680 affected probands with autism, their (largely unaffected) parents, and, when available, their 9,567 unaffected siblings. This study design allows the detection of protein-altering mutations that are present in the proband but not in either parent. These mutations are aggregated by gene and functional category (Satterstrom *et al*., 2026), and the number of observed mutations is compared with its null expectation under the gnomAD mutational model (Karczewski *et al*., 2020). The observed-over-expected number of mutations is an estimate of the *rate ratio*, a measure of effect size. The rate ratio of a mutation is proportional to its penetrance; mutations with a rate ratio of 5 are considered to have “strong” evidence for pathogenicity by American College of Medical Genetics (ACMG) criteria (Richards *et al*., 2015).

The observed number of mutations in gene *g* follows a Poisson distribution whose rate is inflated by the rate ratio:

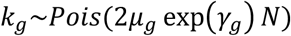

where *k*_*g*_ is the observed count, *μ*_*g*_ is the gene mutation rate estimated using the gnomAD mutational model, and *N* is the sample size such that 2*Nμ*_*g*_ is the expected count under the null, and *γ*_*g*_ is the log-rate ratio. Despite *μ*_*g*_ accounting for trinucleotide context, CpG methylation levels, and depth of sequencing, and being additionally corrected for possible misspecification using synonymous rates in gnomAD (see **Methods**), we emphasize that miscalibration of mutation rates will cause estimation bias. These mutation rate estimates are only available for single-nucleotide substitutions (SNPs), and not for indels, which make up around half of protein truncating variants (PTVs). We exclude frameshift indels from model fitting but include them when estimating the aggregate contribution of *de novo* PTVs, by scaling estimates by the ratio of aggregate PTV count (including frameshift indels) to non-indel PTV count. This approach makes the assumption that frameshift indel functional effects are equivalent to those of protein truncating SNPs in the same gene.

The goal of our study is to infer the distribution of rate ratios *γ*_*g*_ across genes and to characterize several aspects of *de novo* genetic architecture from this inferred distribution. We model the distribution of the log-rate ratio across genes as a mixture of uniform distributions:

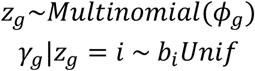

where *z*_*g*_ is a latent variable that assigns genes to mixture components *i*, *Unif* denotes the uniform distribution with extrema at 0 and 1, and *ϕ*_*g*_ denotes the mixture proportions. These mixture proportions are gene-specific because they are allowed to vary as a function of gene-wise annotations (**Methods**); we include loss-of-function observed/expected upper bound fraction, or LOEUF quintile annotations (Karczewski *et al*., 2020), as well as mutation rate, in our model to account for the fact that *de novo* associations with autism are concentrated in large, constrained genes (**Methods**) (Kosmicki *et al*., 2017). We note that such fixed variables that influence mixture proportions are referred to as *concomitant variables* in the statistics literature (Dayton and Macready, 1988).

The approach of fixing a set of mixture components and learning their weights was proposed by Stephens with the *ash* method (Stephens, 2016), and has been used to analyze GWAS effect size distributions (Sinnott-Armstrong *et al*., 2021; Spence *et al*., 2022). It makes the assumption that the distribution has a single mode at zero, which is appropriate because most genes are expected to have little or no effect. We fit the model using sequential quadratic programming (**Methods**), and calculate standard errors and confidence intervals by bootstrapping across genes.

A key estimand is the *mutational variance* (*V*_*μ*_), which is the proportion of variance in binary case-control status that is explained by *de novo* coding mutations aggregated by gene. This quantity is akin to the observed-scale heritability, but *de novo* mutations are not inherited. The mutational variance is the sum of mutational variances across genes:

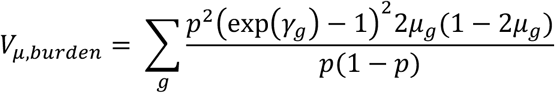

where *p* is the prevalence. This quantity can be computed by integrating over the inferred distribution of gene-wise effect sizes (**Methods**). We note that, because we aggregate variant-wise counts within functional categories (as is standard for this type of study due to the low frequency of individual variants), our estimates are *burden* mutational variances rather than *total* mutational variances. Burden mutational variance is equal to total mutational variance when all variants within a burden mask have equal effect sizes (as is expected for protein-truncating variants), but is less than total mutational variance when variants within a burden mask have different effect sizes (as may be expected for missense variants) (Weiner *et al*., 2023). We mitigate this issue for missense variants by stratifying missense variants into three bins by predicted deleteriousness (Mis0, Mis1, Mis2, with Mis2 predicted to be the most deleterious; see **Methods**). Nonetheless, this stratification of missense variants is likely imperfect, and our estimates of burden mutational variance are likely less than total mutational missense variance. A promising future direction to further address this issue would be inclusion of functional data (such as domain-/residue-level constraint, or protein-protein interaction data) in the design of burden weights to better capture variable variant effects.

A key constant in this equation is the prevalence, which for autism is estimated to be 2.76% in the United States (Maenner *et al*., 2023). For some autism cohorts, cases tend to be more severe, and the phenotype which is ascertained probably has a much lower prevalence. We chose different prevalence estimates for different studies depending on their ascertainment strategy (see **Methods**), estimating the prevalence of autism for the overall study to be 2.2%.

We refer to this method as burdenMLE-DN, or burdenMLE. We evaluated the performance of our method in simulations with varying effect size distributions, which correspond to different patterns of effects across genes. Specifically, we simulated counts using three effect size distributions: “Half-Infinitesimal”, where gene effects are drawn from the positive half of a zero-centered normal distribution, “Point-Normal”, where the majority of gene effects are zero and a small fraction of genes have non-zero effects that are drawn from a normal distribution centered at a positive number, and “Oligogenic”, where the majority of effects are zero and a small fraction of genes have a large non-zero effect (**Figure 1A**). These simulations demonstrate that burdenMLE produces well-calibrated parameter estimates at N > 1000 probands (**Figure 1B**). We found that at current sample size, estimates of mutational variance were approximately unbiased for each of these distributions, with subtle downward bias for oligogenic architectures, likely due to the fact that they differ most sharply from unimodality (**Figure 1B**). We additionally evaluated estimation unbiasedness for other quantities from our analyses, and found that they were similarly approximately unbiased across varying effect size distributions at current sample sizes (**Supplementary Figure 1).** Simulation results are available in **Supplementary Table 1**.

**Figure 1.**
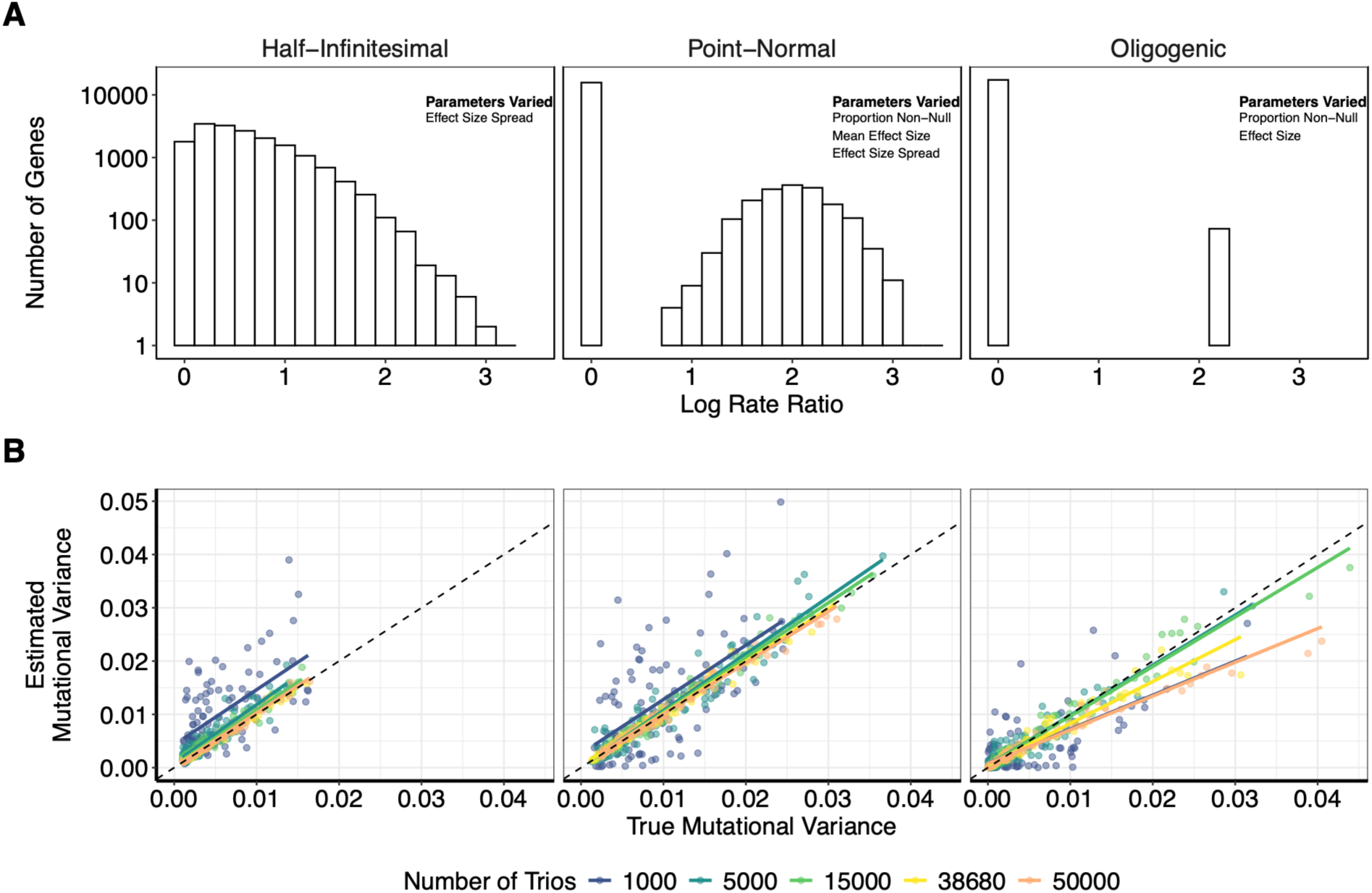
Simulations. **(A)** Sample histograms for infinitesimal, point-normal, and oligogenic effect size distributions, the three effect size distributions modelled in our simulations. These distributions reflect the cases where effect sizes are drawn from a single half-normal component (Half-Infinitesimal), when most effects are null and a small number of effects are non-null drawn from a positive-centered normal distribution (Point-Normal), and when most effects are null with a small number of non-null effects with a large positive effect (Oligogenic). Text in the upper right of each plot describes which parameters are varied across simulations. For each simulation, we drew gene-wise effect sizes from one of these distributions, simulated mutation counts at varying sample sizes using a Poisson model, and analyzed the resulting counts with burdenMLE. **(B)** Comparison of true and estimated mutational variance (i.e. observed-scale phenotypic variance explained by the burden of *de novo* coding variants) values for varying effect size distributions and sample sizes. Each point represents one simulation with a particular choice of parameters described in Panel A.

### Population and clinical genetic characterization of de novo contributions to autism

We analyzed a new dataset from the Autism Sequencing Consortium (ASC), which processed and aggregated exome-sequencing data from multiple sources (Satterstrom *et al*., 2026). Their combined sample size was 38,680 affected proband trios, spread across three overarching cohorts: Simons Powering Autism Research (SPARK; N = 18060), the Autism Sequencing Consortium (ASC; N = 8679, including 2422 from the Simons Simplex Collection), GeneDx, a clinical testing company (N = 10747) (**Figure 2A, Supplementary Table 2**; **Methods**), as well as a smaller cohort of individuals with autism from the Deciphering Developmental Disorders cohort (DDD; N = 1194). We stratified the 47933 observed *de novo* coding variants into 6470 protein truncating variants (PTVs, of which 3077 were frameshift indels), 30110 missense variants (of which 24405 were Mis0, 3443 were Mis1, and 2262 were Mis2; **Methods**), and 11353 synonymous variants (Satterstrom *et al*., 2026). For each of these variant categories, we estimated the distribution of gene-wise burden effect sizes, and computed the observed-scale coding mutational variance from these inferred distributions (**Methods; Supplementary Table 3**). All of our analyses are restricted to autosomal mutations.

**Figure 2.**
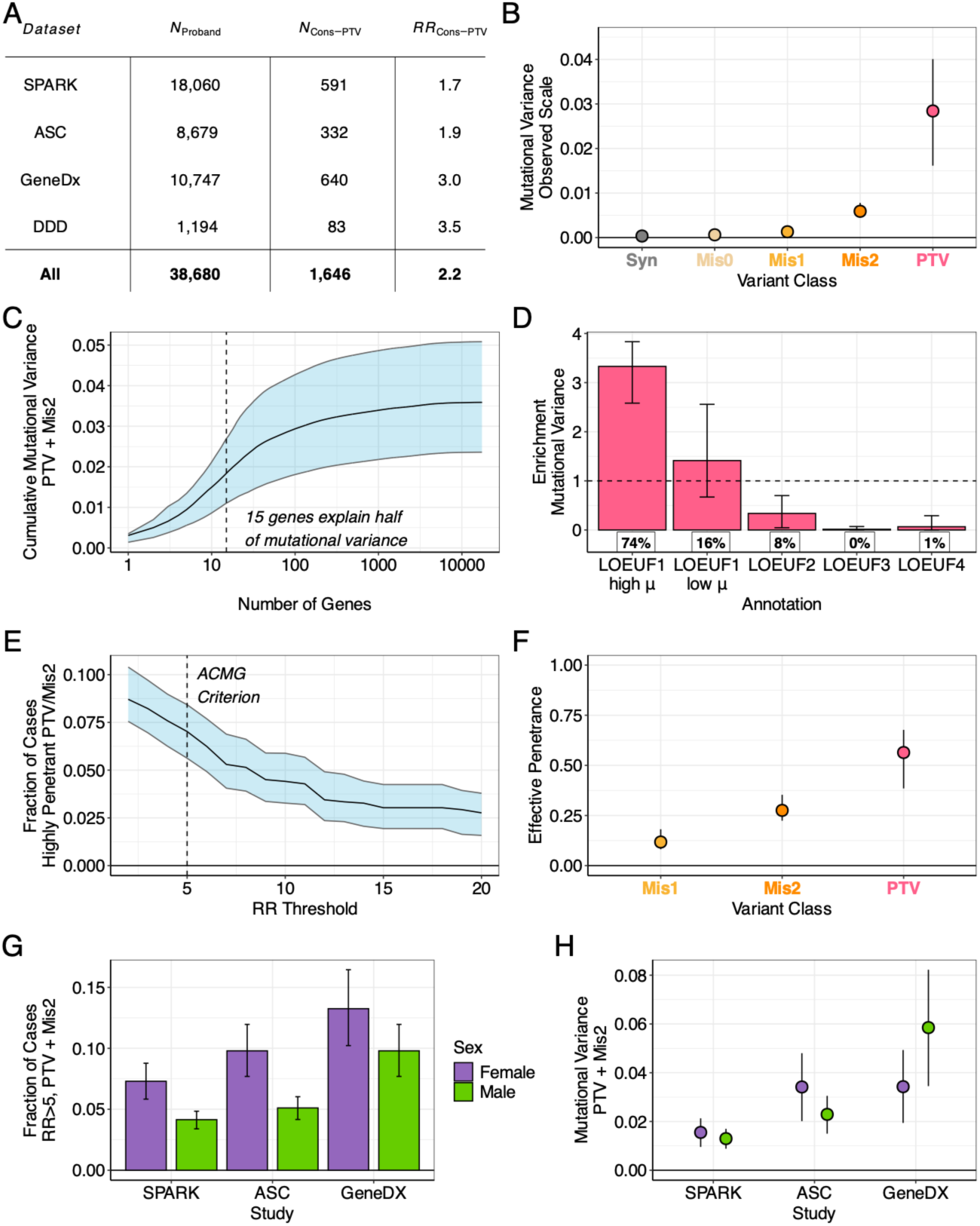
**(A)** Description of component datasets. Rate ratio in constrained PTVs was computed by dividing the number of PTVs in the top quintile of genes by LOEUF by the sum of gene-wise mutation rates for this set of genes multiplied by 2*N*. **(B)** Mutational variance estimates of autism in the aggregated dataset for five variant classes. **(C)** Fraction of mutational variance captured by differing number of genes. Shaded error bar represents 95% confidence interval. Vertical dashed line is drawn at the number of genes needed to explain 50% of mutational variance. **(D)** Enrichment of mutational variance in gene annotations. Error bars indicate 95% confidence interval. Horizontal dashed line indicates null enrichment value of 1. Fraction of mutational variance in each gene set is shown below the bars. **(E)** Fraction of cases carrying a large-effect PTV/Mis2 variant at various effect size thresholds. Shaded error bar represents 95% confidence interval. Vertical dashed line is drawn at the ACMG “strong” pathogenicity evidence threshold of 5. **(F)** Estimates of effective penetrance for three variant classes. Error bars represent 95% confidence intervals. **(G)** Fraction of cases carrying PTV/Mis2 variant with rate ratio of at least 5 in the three component datasets, stratified by sex. Error bars represent 95% confidence intervals. **(H)** Mutational variance estimates aggregated between PTV and Mis2 in the three component datasets, stratified by sex. Error bars represent 95% confidence intervals.

The observed-scale coding mutational variance of autism was estimated to be 2.8% for PTVs (95% CI: 1.6% - 4.0%), 0.6% for Mis2 variants (95% CI: 0.5% - 0.8%), 0.1% for Mis1 variants (95% CI: 0.08% - 0.2%), 0.06% for Mis0 variants (95% CI: 0.03% - 0.10%), and 0.04% for synonymous variants (95% CI: 0.01% - 0.08%) (**Figure 2B**). Aggregating between PTVs and Mis2 variants, we estimate that the total observed-scale autosomal coding mutational variance of autism is approximately 3.4% (95% CI: 2.1% - 4.7%). We observed near-zero mutational variance (0.013%) in unaffected siblings (**Supplementary Figure 2**).

We next proceeded to assess the sensitivity of these estimates to various factors. There was little effect when we removed 69 genes with evidence of clonal expansion in spermatogonia, which is a potential confounder in trio-based association studies (Neville *et al*., 2025; Seplyarskiy *et al*., 2025) (**Supplementary Figure 3**). Additionally, we compared paternal ages in the SPARK cohort to paternal age estimates for the general US population from the CDC, finding only a modestly higher mean paternal age in the SPARK cohort (∼5 months, Cohen’s d = 0.06; **Supplementary Figure 4**), which is expected to cause a negligible upward bias of 0.019% in combined PTV/Mis2 mutational variance (**Methods**). These estimates vary across different choices of prevalence (**Supplementary Figure 5)**; for example, using a prevalence of 1% (which would be appropriate if ascertained cases are more severe than the general autism population), the mutational variance aggregating PTVs and Mis2 variants is 1.5% (95% CI: 0.9% - 2.1%). To assess agreement with a commonly used Bayesian association framework (TADA, Transmission and *De Novo* Association) (He et al, 2013), we computed posterior-mean rate ratios using BurdenMLE and compared them with TADA-derived Bayes factors from the most recent ASC analysis (Satterstrom *et al*., 2026),and found that these quantities were positively correlated (**Supplementary Figure 6**). To evaluate application of BurdenMLE in a smaller and ancestrally distinct dataset, we analyzed 680 affected offspring from a Korean autism whole-genome sequencing study (Kim *et al*., 2024), finding that mutational-variance estimates were imprecise at this sample size (PTV: 0.09%, 95% CI 0.04%–4.43; pooled missense: 2.22%, 95% CI 0.44%–5.7%) (**Supplementary Figure 7**).

Across complex traits, rare and common variants differ in their polygenicity, and rare-variant burden heritability is much more strongly concentrated in a smaller number of genes (Weiner *et al*., 2023). For autism, its common variant architecture is extremely polygenic (Grove *et al*., 2019); we quantified the degree of polygenicity for *de novo* coding mutations (**Figure 2C; Methods**). We found that 50% of the autosomal mutational variance is contained in 15 genes, with 95% of the mutational variance contained in approximately 1100 genes (**Supplementary Table 4)**. We note that these 15 genes are also strongly associated with severe developmental disorders (Kaplanis *et al*., 2020). In contrast, no locus explains an appreciable fraction of the common variant heritability of autism (Grove *et al*., 2019). For example, *SCN2A* alone explained 8.6% of the mutational variance of autism in the current samples.

In the general population, PTVs are strongly depleted in genes associated with rare variant association to autism (Kosmicki *et al*., 2017). We quantified the enrichment of mutational variance in loss-of-function constrained genes (Karczewski *et al*., 2020) and found that the top 20% of constrained genes explain 90% of mutational variance (95% CI: 81% - 97%); all other bins were significantly depleted (**Figure 2D**). The enrichment was driven entirely by the subset of constrained genes that were also in the top 20% by mutation rate (i.e., long genes). This correlation between effect size and mutation rate may be due to the two groups of genes most strongly associated with autism, gene expression regulation and neuronal communication (Satterstrom *et al*., 2020), which have median coding sequence length 1.2x and 1.3x the median of all other genes, respectively (**Supplementary Figure 8**). Indeed, we estimated mean coding sequence length at varying rate ratio thresholds, finding that the mean coding sequence length is greater for larger-effect genes (**Methods**; **Supplementary Figure 9**). Importantly, our analysis accounts for gene size by modelling variation in gene-wise mutation rate; this is unlike significant gene discovery, where larger genes have greater statistical power due to their larger mutation rates. Our results imply that, in addition to having larger mutation rates, large constrained genes also tend to have larger effect sizes for autism.

If all autism-causing mutations were fully penetrant, then the mutational variance would equal the fraction of cases carrying a causal allele, but empirically, many mutations associated with autism have effect sizes that suggest incomplete penetrance (Neale *et al*., 2012; Fu *et al*., 2022). We estimated the fraction of cases with a large-effect mutation at varying rate-ratio thresholds, integrating over effect size posterior distributions for each gene (**Figure 2E; Methods; Supplementary Table 5**). We estimate that 7.0% of autistic individuals in our cohorts carry a PTV or Mis2 variant with rate ratio greater than 5 (95% CI: 5.6% - 8.4%), which has been suggested by the American College of Medical Genetics as “strong” evidence of variant pathogenicity (Richards *et al*., 2015). With a prevalence of 2.2%, this rate ratio corresponds to a diagnosis probability (penetrance) of 0.11. At a more stringent threshold of RR = 20 (penetrance = 0.44), the fraction of carriers decreases to 2.8% (95% CI: 1.6% - 3.8%). More generally, there exists a wide penetrance spectrum across genes.

How can the penetrance spectrum be summarized? We defined the *effective penetrance* to be the average penetrance of an excess mutation in cases relative to the null mutational mode (**Figure 2F; Methods; Supplementary Table 3**). For example, in a non-causal gene, there are no excess mutations, and it does not contribute to the average penetrance. We estimate that the effective penetrance is 0.56 for PTVs (95% CI: 0.38 – 0.68), 0.28 (95% CI: 0.22 – 0.35) for Mis2, and 0.12 for Mis1 (95% CI: 0.08 – 0.18). For PTVs, this estimate suggests that the average causal *de novo* mutation for autism is associated with a ∼50% chance of diagnosis. Our estimates suggest that the average autism-associated *de novo* variant is indeed highly penetrant, but certainly not fully penetrant. These estimates can also be expressed on the rate ratio scale; the *effective rate ratio* is 25 for PTVs (95% CI: 17 – 31), 12 for Mis2 (95% CI: 10 - 16), and 5.3 for Mis1 (95% CI: 3.7 – 8.2). We caution that for a complex phenotype such as autism, incomplete penetrance may reflect diagnostic uncertainty in addition to variability of effects. This would tend to deflate penetrance estimates if some carriers are affected but do not receive diagnoses, even with clinically significant neurodevelopmental phenotypes.

Autism is a highly heterogeneous phenotype (Lord, 2019), which may lead to differences in effect size estimates across strata. The three cohorts in our dataset vary widely in their ascertainment strategy, in a manner that is reflected in molecular data: GeneDx samples, which come from autistic individuals referred for clinical genetic testing, had a much larger excess of *de novo* PTVs in loss-of-function constrained genes than samples from SPARK or ASC, which have broader ascertainment (**Figure 2A**). Consequently, GeneDx samples additionally have greater mutational variance and fraction of cases with large effect (RR>5) variants (**Supplementary Figure 10**). Another key axis of heterogeneity is sex, where female autistic individuals carry a greater burden of rare *de novo* mutations (Fu *et al*., 2022). Accordingly, we estimate that a larger fraction of female probands are carriers of large-effect (RR>5) variants (**Figure 2G; Methods; Supplementary Table 6**). In contrast, our estimates of mutational variance are much more stable across sex (**Figure 2H; Methods; Supplementary Table 6**). These smaller differences in mutational variances are consistent with the opposing actions of gene-wise effect sizes (which are larger in females and increase female mutational variance; **Supplementary Table 2**) and prevalence (which is lower in females and decreases mutational variance) (**Methods**). Lastly, we considered the role of developmental delay and/or intellectual disability (DD/ID), which is a frequent comorbidity in autism (Maenner *et al*., 2023) and known to influence the genetic architecture of autism (Robinson *et al*., 2014). We note that this analysis is affected by variability of DD/ID definition across cohorts due to differing availability of phenotypic information (**Methods**). We found that, as expected, comorbid DD/ID was associated with increased mutational variance, increased fraction of probands carrying a large-effect (RR>5) variant, and increased effective rate ratio of excess mutations (**Supplementary Figure 11**). To assess the patterning of genetic architecture across all of these proband variables, we fit models that simultaneously stratify by cohort, sex, and DD/ID status (**Supplementary Figure 12**). The resulting parameter estimates are difficult to interpret in detail due to low power in these fine-grained strata.

### Future autism gene discovery

To date, trio-based exome sequencing studies have identified hundreds of genes with significant excess of *de novo* loss-of-function variants in autism (Satterstrom *et al*., 2026). As sample sizes reach the tens of thousands, this raises the question of how many genes remain to be found, and the magnitude of their effect sizes. Genes may be yet undiscovered because of incomplete penetrance (Zhou *et al*., 2022) or low mutation rate, which determines statistical power.

From our inferred *de novo* effect size distributions, we estimated the number of autism-associated genes at various effect size thresholds (**Figure 3A**). We estimate that there are 601 genes with rate ratio greater than 2 (95% CI: 422-838); this estimate is roughly consistent with, but generally lower than, previous estimates of the number of autism-associated genes (119 - 2555 in (Sanders *et al*., 2012), less than 1000 in (Neale *et al*., 2012), 550 - 1000 in (He *et al*., 2013)). Using our more detailed model of genetic architecture, we also estimated how many genes exist with larger effect sizes. Approximately half of autosomal autism-associated genes (312; 95% CI: 232-362) have rate ratio greater than 5, and a smaller fraction have larger effect sizes (124 or 21% with RR > 10; 95% CI: 88-167, 46 or 8% with RR > 20; 95% CI: 28-62).

**Figure 3.**
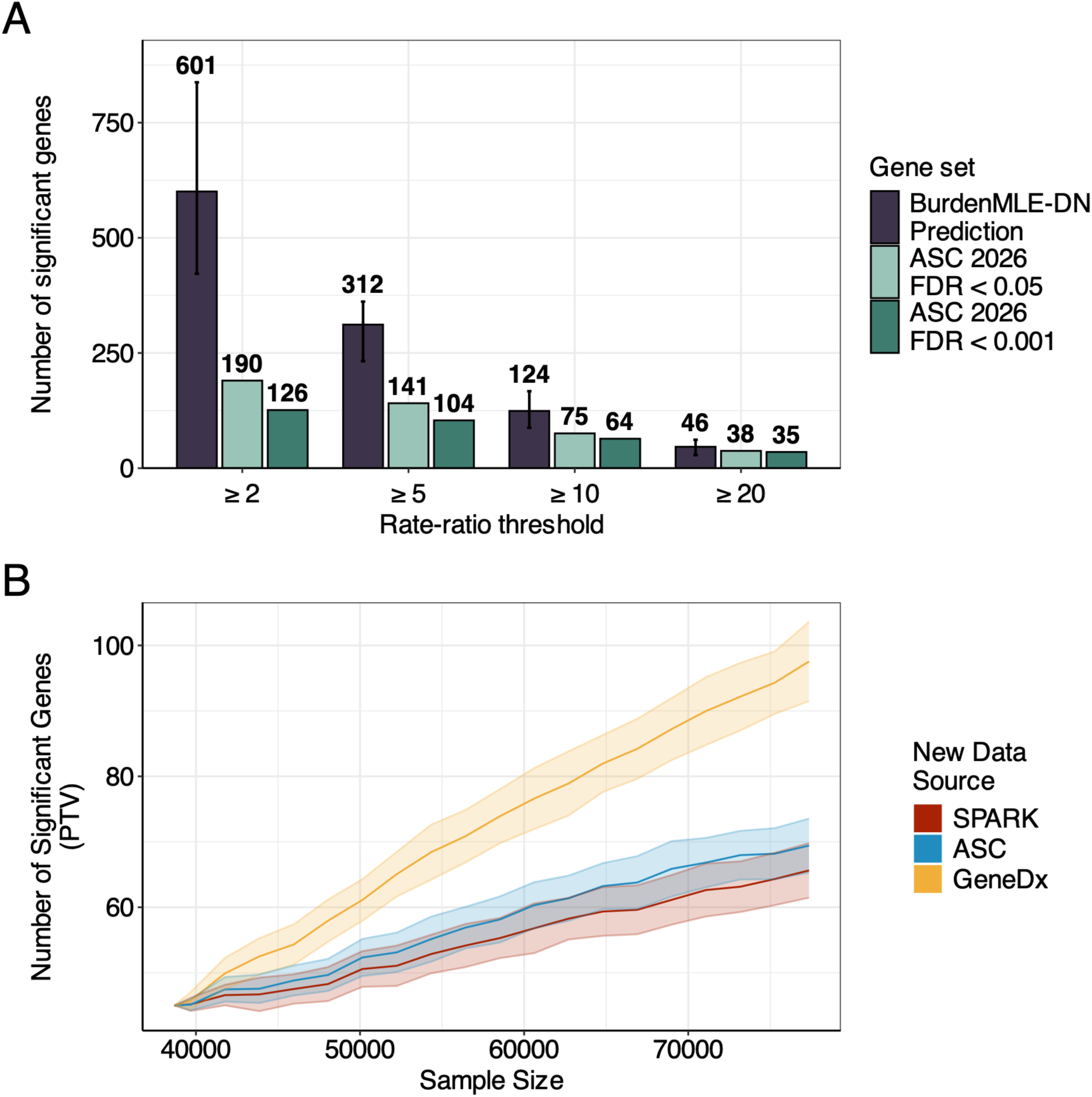
Forecasting gene discovery. **(A)** Bar chart with the estimated number of genes with varying minimum effect size, and the number of previously discovered genes with varying minimum effect size, using gene sets from Satterstrom et al, 2026. (**Methods**). **(B)** Predicted gene discovery via PTV associations at future sample sizes, color represents the source of the new data. Shaded error bars represent one standard deviation.

We then compared these estimates with the most recent published gene discovery effort from the Autism Sequencing Consortium from which our data are derived (Satterstrom *et al*., 2026). We found that across the penetrance spectrum, many more genes remain to be discovered (**Figure 3A**). In particular, we estimate that current *bona fide* autism-associated genes (FDR < 0.001) represent 21% of the total number of autism-associated genes with rate ratio greater than 2 (126 discovered out of estimated 601); this percentage rises to 32% when using a more liberal significance threshold (FDR < 0.05). A larger proportion of highly penetrant genes have already been discovered (for rate ratio greater than 20, 35 of estimated 46 have been discovered), but, nonetheless, approximately one quarter of highly penetrant genes remain to be found.

Based on our previous findings related to cohort heterogeneity, we hypothesized that the pace of gene discovery would vary substantially depending on the source of new data. We estimated gene discovery trajectories, assuming that new data comes from the SPARK, ASC, and GeneDX cohorts (**Methods; Supplementary Table 7**). We specifically estimated gene discovery from *de novo* PTVs; this differs from the standard tool for ASD gene discovery (TADA) (He *et al*., 2013; Fu *et al*., 2022), which integrates evidence across many variant and study types, but is difficult to formally analyze. We found that the relationship between sample size and number of discovered genes varied dramatically with ascertainment of new samples (**Figure 3B**). When we simulated new data being drawn from the GeneDx cohort, new genes were discovered much more quickly, owing to the greater excess of *de novo* mutations in this cohort; the number of new discoveries was lower in the SPARK and ASC cohorts. This finding implies that strategic ascertainment may accelerate gene discovery efforts (Peloso *et al*., 2016), but also raises questions about whether discovered genes may be specific to particular cohorts, such as the GeneDx cohort, which has a higher rate of comorbid intellectual disability. Indeed, when we conducted a similar analysis forecasting gene discovery when new samples are drawn from probands with or without comorbid DD/ID, we found that the pace of gene discovery is much faster if new probands have comorbid DD/ID (**Supplementary Figure 13**).

### Comparison with severe developmental disorders

*De novo* mutations are strongly associated with a wide range of severe developmental disorders, including growth abnormalities, birth defects, and disorders of learning and behavior (Deciphering Developmental Disorders Study, 2017). The symptoms of developmental disorders can include a diagnosis of autism, and developmental-disorder-associated genes overlap with autism-associated-genes (Kaplanis *et al*., 2020; Satterstrom *et al*., 2020). It has been previously estimated that 13.4% and 28.4% of probands carry diagnostic *de novo* PTV and missense variants, respectively (Deciphering Developmental Disorders Study, 2017), and that roughly 1,000 undiscovered genes contribute to developmental disorders with PTV rate ratios of around 10 (Kaplanis *et al*., 2020).

We applied burdenMLE to estimate the distribution of *de novo* burden effect sizes for developmental disorders, using the 31,058 trios from Kaplanis et al (2020). We estimated that the observed scale mutational variance of developmental disorders is 6.5% for PTVs (95% CI: 4.1% - 8.6%), 1.7% for Mis2 variants (95% CI: 1.3% - 2.2%), 0.5% for Mis1 variants (95% CI: 0.3% - 0.6%), 0.1% for Mis0 variants (95% CI: 0.09% - 0.17%), and 0.02% for synonymous variants (95% CI: 0.02% - 0.03%) (**Figure 4A; Supplementary Table 8**). Aggregating across PTVs and Mis2 variants, we estimate that the mutational variance of developmental disorders is 8.3% (95% CI: 5.9% - 10.4%), approximately 2.4x larger than that of autism, although these estimates are sensitive to the assumed prevalence (1% for developmental disorders; **Methods**; see **Supplementary Figure 14** for estimates across a plausible range of prevalence values). We estimated the fraction of DD cases carrying a large-effect variant at various rate ratio thresholds, finding that 16.3% of developmental disorder probands carry a PTV or Mis2 variant with rate ratio greater than 5 (95% CI: 14.0% - 18.6%) versus 7.0% for autism (95% CI: 5.6% - 8.4%) (**Figure 4B; Supplementary Table 9**). We also estimated the effective rate ratio (i.e. the average rate ratio of excess mutations; **Methods**), which was 60 for PTVs (95% CI: 46-77; **Figure 4C**); this number is higher than in autism (25 for PTVs, 95% CI: 17-31; **Figure 2F**) but similar in units of penetrance due to the difference in prevalence. Developmental disorders are known to be polygenic; we estimated that 341 genes harbor PTVs with a rate ratio of at least 5 (95% CI: 259 - 379), similar to autism (312; 95% CI: 232-362) (**Figure 4D**). Our analysis retains 3543 overlapping samples in both cohorts; when we remove these samples from the autism dataset and retain them in the DD dataset, we obtained a similar pattern of results (**Supplementary Figure 15**).

**Figure 4.**
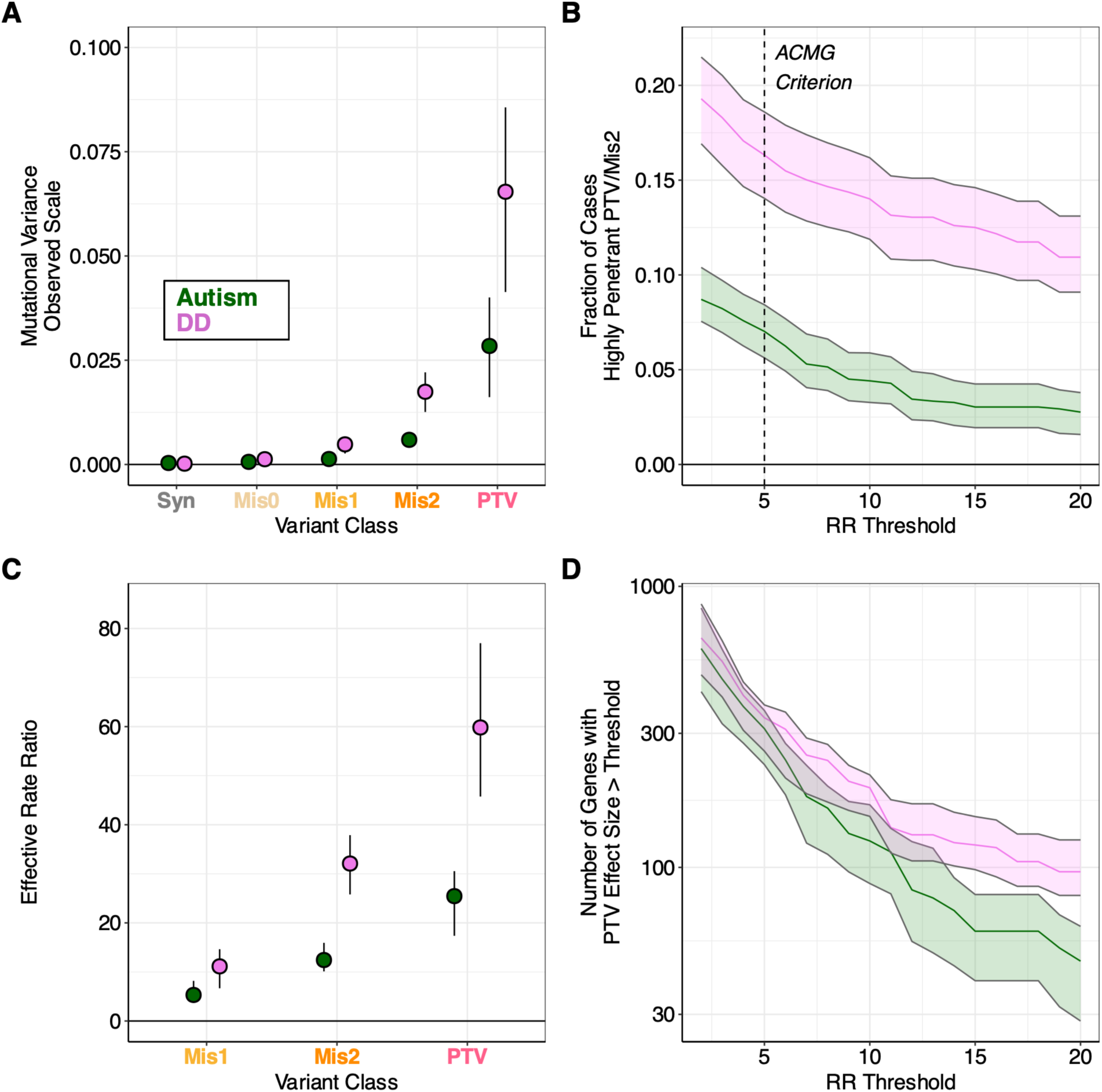
Comparison with developmental disorders. **(A)** Comparison of mutational variance, error bars indicate 95% confidence interval. **(B)** Comparison of fraction of cases carrying a large-effect PTV/Mis2 variant at various effect size thresholds. Shaded error bar represents 95% confidence interval. Vertical dashed line is drawn at the ACMG “strong” pathogenicity evidence threshold of 5. **(C)** Comparison of effective rate ratio estimates, error bars represent 95% confidence intervals. **(D)** Estimates of the number of genes with various minimum effect sizes, error bars represent 95% confidence intervals.

Compared with previous estimates, our estimates suggest a more modest contribution of *de novo* coding mutations to developmental disorders. In particular, our estimate for the mutational variance (8.3%) is smaller than the reported excess of *de novo* mutations (42.2 per 100 cases, Deciphering Developmental Disorders Study, 2017); these numbers would be expected to agree if all mutations were either fully penetrant or null. We used different variant annotations and mutation rates as compared with Kaplanis et al (2020), but we still find a similar excess of mutations (10.0 excess PTVs and 25.9 excess missense mutations per 100 cases; **Methods**). Instead, the discrepancy is explained by incomplete penetrance: for example, for a condition with 1% prevalence, if 100% of cases carry a pathogenic mutation but only 10% of carriers are cases, then the excess mutation rate is 90 mutations per 100 cases, but the mutational variance is only 9%. This effect is especially strong for Mis0 and Mis1 mutations, which have a low effective rate ratio but contribute significantly to the mutational excess (18 per 100 cases, vs. 0.6% mutational variance). For PTVs, with their high effective rate ratio, the discrepancy is smaller (10 per 100 cases vs. 6.5%).

## Discussion

In our analyses, we have estimated several features of the *de novo* genetic architecture of autism. We estimate that the autosomal mutational variance of autism due to damaging SNPs and frameshift indels is 3.4%; that the fraction of cases carrying a “large-effect” (RR>5) *de novo* coding variant is 7.0%; that a typical autism-causing protein-truncating variant is 56% penetrant; that these features vary substantially across cohorts; and that there are many more autism-associated genes to be found across the effect size spectrum.

Like all analyses of autism, these estimates are contingent on a particular phenotypic definition and ascertainment strategy, which can vary substantially (Kuo *et al*., 2022). We characterized heterogeneity across cohorts that vary in ascertainment, finding differences in the fraction of cases with large-effect variants (**Figure 2G**). Consequently, the yield in gene discovery is greater per sample in the GeneDx cohort, where probands are referred for clinical genetic testing and generally have more severe phenotypes than in the other cohorts **(Figure 3B**). Our estimates of the mutational variance are contingent upon assumed values of the cohort-specific prevalence, and more generally upon an assumed model of ascertainment (**Methods**). If the prevalence were overestimated (as may be the case for more severely affected cohorts), it would upwardly bias our estimates of the mutational variance, and vice versa (**Supplementary Figure 5**). Similarly, if SPARK and ASC exhibit cohort ascertainment bias relative to the general autistic population, it would cause bias, possibly in either direction, in our estimates of the mutational variance and the effective penetrance.

In combination with similar analyses of common genetic variation (Yang *et al*., 2010; O’Connor, 2021) and segregating rare variation (Wainschtein *et al*., 2022; Weiner *et al*., 2023), our analyses of *de novo* variants present an opportunity to reflect on the patterning of genetic associations across the entire allele frequency spectrum. Firstly, we estimate that the vast majority of mutational variance for autism resides in loss-of-function constrained genes (**Figure 2D**). This concentration is greater than that which we previously observed in either segregating rare coding variants or common variants (Weiner *et al*., 2023). This trend is consistent with large-effect genes being highly constrained, and also making a greater relative contribution to variance at lower allele frequencies. Conversely, common variant associations may implicate either large or small effect genes, and are therefore expected to be much more polygenic (O’Connor *et al*., 2019). Indeed, we observe that for autism, mutational variance is explained by a small number of genes, contrasting sharply with its common-variant architecture.

Although mutational variance is unknown for most human traits and diseases, mutational variance has been estimated for many traits in other organisms in the population genetics literature by analyzing mutation accumulation experiments (Houle, Morikawa and Lynch, 1996). Our estimated mutational variance of autism is larger than that of all traits compiled in Lynch & Walsh (Lynch and Walsh, 1998), which is especially notable given that our estimates are not inclusive of copy-number and non-coding variation. Most likely, it is also larger than most common conditions in humans; in particular, a trio study of schizophrenia (n=2541 trios) found inconsistent evidence for enrichment of deleterious *de novo* mutations in affected probands (Howrigan *et al*., 2020). The mutational variance of a phenotype at equilibrium is closely connected with strength of selection (Lynch and Walsh, 1998), consistent with the known impact of autism on fecundity (Power *et al*., 2013).

Several of our analyses are relevant to the clinical genetics of autism. Using the ACMG “strong” pathogenicity criterion of rate ratio = 5 as an arbitrary cutoff, we estimate that 7.0% of autism cases carry a damaging *de novo* coding mutation, inclusive of SNPs and frameshift indels (**Figure 2E**). We emphasize that because our model does not include copy-number variants, non-coding variants, or recessive effects, our estimates do not capture the entire diagnostic yield. We also estimate that autism-associated *de novo* variants have an average penetrance of 0.56, 0.28, and 0.12 for PTVs, damaging missense variants (Mis2), and less damaging missense variants (Mis1), respectively (**Figure 2F**). These estimates suggest that, for the classes of genetic variation considered, trio exome sequencing will provide genetic diagnoses for a minority of autistic individuals. Most of these variants are incompletely penetrant, similar to other conditions (C. F. Wright *et al*., 2024), but still exceed the ACMG criterion of “strong” evidence of pathogenicity (penetrance = 0.138, assuming prevalence of 2.76%) (Maenner *et al*., 2023). We note, however, that penetrance in our model refers simply to the phenomenon that not all carriers of a variant in a given burden mask have a diagnosis of autism. This could be due to several factors, including (1) inclusion of null mutations in a burden mask, i.e. poorly annotated missense variants, (2) variability in background risks, where some probands have lower or higher familial diathesis that change whether a *de novo* mutation raises liability beyond a diagnosis threshold, (3) diagnostic complexity, where for example some variants may sometimes cause co-morbid severe epilepsy or ID, where autism may or may not be diagnosed, and (4) genuine stochasticity of mutational effects, where the biological consequences of a mutation are variable. In future work, it may be possible to disambiguate these factors by embedding burdenMLE in a larger model that incorporates detailed functional variant annotations, familial risk, rich phenotypic and comorbidity data, and biomarkers of biological effects.

Our forecasting analyses clarify the expected yield of increased investment in exome-sequencing of autistic individuals. We find that not only are many autism genes yet undiscovered in genome-wide analyses, but that many undiscovered genes have very large effect sizes (**Figure 3**). We note that these findings do not account for important contributions to gene discovery from other approaches, such as sequencing of related developmental disorders and case series. Our findings suggest that mutation rate, rather than effect size, is often the primary limiting factor in gene discovery, and that increasing sample size will continue to identify highly penetrant genes. On the other hand, our finding that the mutational variance of autism is strongly concentrated in a small number of genes (with 50% in only 15 genes; **Figure 2C**) suggests that these genes will make only modest contributions to genetic diagnosis. This is explained by the fact that low mutation rates limit both diagnostic yield and statistical power for gene discovery. With this in mind, we posit that as sample sizes increase, the primary utility of gene discovery will shift from diagnostic yield (with discovery of genes that explain a large fraction of cases) to biological insight (with discovery of genes across the effect size spectrum that explain a small number of cases). This does not dampen our enthusiasm for investment in increased sample size, as we expect that the biological insight conferred by a newly discovered gene is largely unrelated to its mutational frequency in cases.

## Methods

### Overview of methods

In a *de novo* association study, we use a trio design to identify variants that are present in offspring but not in parents. When the observed number of *de novo* variants in a gene exceeds the expected number of *de novo* variants given a null mutational model, we infer that variants in that gene are associated with the phenotype of interest. In the studies we consider, we focus specifically on coding variants identified via exome sequencing. We parameterize effect sizes using a Poisson model:

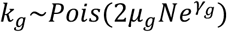

Where *μ*_*g*_ is the cumulative gene mutation rate, *N* is the number of trios in the analysis, and *γ*_*g*_is the log rate ratio. Under this model, the effect sizes *γ*_*g*_ describe the multiplicative change in the frequency of *de novo* variants in gene *g* relative to the expectation under a null mutational model. We aim to define and estimate the distribution of *de novo* burden effect sizes over genes. We do so via a hierarchical model, where gene-wise effect sizes *γ*_*g*_ arise from a mixture of uniform distributions:

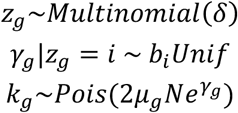

Where *b*_*i*_ is the mixture component parameter (i.e. the extremum of the uniform mixture component), *Unif* denotes the *Uniform*(0,1) distribution, *z*_*g*_ encodes the mixture component that gene *g* belongs to, and *δ* are the mixture proportions.

We pre-select a grid of uniform mixture component extrema *b*_*i*_ ranging from 0 to a positive number to make inference tractable (see *burdenMLE implementation details* below). One of these mixture components is *Unif*(0,0), corresponding to a point mass at zero. There are no mixture components with negative point mass, corresponding to a choice not to model protective effects; we elected to do so due to the low power to detect protective effects. Our usage of uniform mixture components to model an effect size distribution closely follows Stephens (Stephens, 2016), who used a similar model to shrink effect sizes. Combinations of these mixture components span any unimodal distribution with mode at zero.

The estimand of burdenMLE is *δ*, which, in conjunction with mixture component parameters *b*_*i*_, defines the distribution of effect sizes *γ*_*g*_ across genes.

### Model fitting: Sequential quadratic programming

We estimated mixture weights using the mixsqp R package (version 0.3.54), which uses sequential quadratic programming to maximize the likelihood of a finite mixture with fixed component distributions (Kim *et al*., 2018). Briefly, mix-SQP locally approximates the convex mixture-likelihood objective with a quadratic function and iteratively solves the resulting simplex-constrained quadratic subproblems. An active-set algorithm efficiently solves these subproblems by concentrating computation on mixture components relevant to the current solution.

Models were initialized with uniform mixture weights. After finding that the built-in EM warm start led to early elimination of low-probability mixture components, we disabled warm-start (numiter.em = 0) and prevented small fitted weights from being automatically set to zero (zero.threshold.solution = 0). Each bootstrap replicate was likewise initialized with uniform weights, ensuring that all components were available to the optimizer. To accommodate models with different numbers of components, we set the maximum number of active-set iterations per quadratic subproblem to Max(20,2*K*) where *K* is the number of mixture components. We confirmed in simulations that mixsqp produced similar estimates to the expectation-maximization algorithm described below, with much faster runtime (**Supplementary Figure 16**).

### Model fitting: Expectation-maximization algorithm

We also developed a tool to estimate *δ* with expectation maximization, which has been extensively studied in the setting of finite mixture models (Hastie, Tibshirani and Friedman, 2009). The procedure alternates between computing the posterior probabilities of component membership (E-step) and maximizing the expected complete-data log-likelihood with respect to *δ* (M-step).We first initiate some values of *δ*.

#### E-step

In the E-step, we compute the posterior probability of component membership for each gene, or *responsibility,* conditioning on *δ*^(*t*)^, the current mixture coefficients. The responsibility of component *i* for gene *g* is:

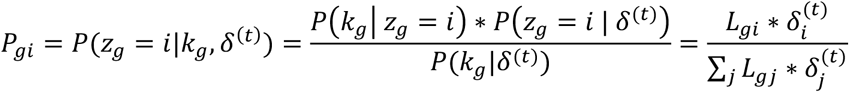

The *conditional likelihood L*_*gi*_ can be efficiently precomputed for each gene/component pair as

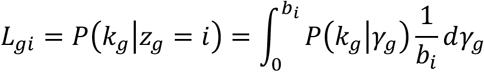

Where 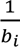 is the *b Unif* density. The distribution implied by this likelihood is a compound Poisson-Uniform distribution. We numerically approximate this integral by calculating the average conditional likelihood across several values of *γ*_*g*_ along the domain of *γ*_*g*_ | *z*_*g*_ = *i* ∼ *b*_*i*_*Unif*.

#### M-step

In the M-step, we seek to find the value of *δ* that maximizes the expected complete log-likelihood:

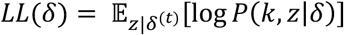

Expanding, and removing constants:

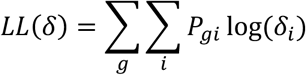

We seek to maximize this quantity with respect to *δ* subject to the constraint that ∑_*i*_ *δ*_*i*_ = 1. Introducing a Lagrange multiplier *λ* for the constraint:

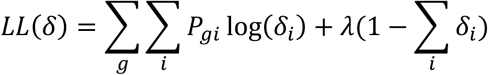

Taking partial derivatives with respect to *δ* and setting 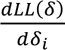 equal to 0 yields the standard update equation for mixture proportions *δ*:

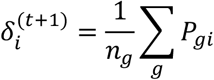

I.e. the average of the responsibilities for that component across genes; this algorithm has previously been derived in detail (Hastie, Tibshirani and Friedman, 2009). We repeat the E- and M-steps until the proportional change in *LL*(*δ*) is less than a user-specified tolerance parameter.

### burdenMLE implementation details

#### Choice of mixture components

We fit models with ten half-uniform mixture components, with extrema spaced evenly between 0 (i.e. a point-mass at zero) and log(100). The maximum of log(100) assumes that the prevalence of autism is no less than 1%, in which case a rate-ratio of 100 would imply full penetrance. This choice is based on recent analyses suggesting that 1% is a lower-bound prevalence estimate for severe developmental disorders (Seplyarskiy *et al*., 2025). We did not include components ranging over negative values (i.e. protective variants) due to the very low power to detect protective associations in current datasets.

#### Poisson-Uniform evaluation grid

To evaluate Poisson-Uniform likelihoods as described above, we use a grid size of 10, i.e. we average over 10 evenly spaced values across the range of the mixture components *b*_*i*_*Unif*.

#### Expectation-maximization convergence parameter

We run the expectation-maximization algorithm until the proportional change in log likelihood is less than 1e-6,with a maximum number of iterations of 10000.

#### Mutation rates

We used mutation rates from gnomAD v4 (Chen *et al*., 2024), which we corrected using burdenMLE, see *Mutation Rate Misspecification* below

#### Variant stratification

We stratified coding variants into five variant classes: protein-truncating variants (PTVs), three classes of missense variants (Mis0, Mis1, Mis2), and synonymous. PTVs were identified using LOFTEE (Karczewski *et al*., 2020), retaining variants that were either labelled as high-confidence loss-of-function variants or “other splice” (OS) variants. OS variants were included based on Autism Sequencing Consortium analyses suggesting that OS variants have similar effect sizes to high confidence loss-of-function variants. Missense variants were stratified via two filters: Missense deleteriousness Prediction by Constraint (MPC) > 2 (Chao *et al*., 2024) and AlphaMissense pathogenicity (AM) > 0.97 (Cheng *et al*., 2023). Mis0 variants met neither of these criteria, Mis1 variants met one of these criteria, and Mis2 variants met both of the criteria. Synonymous variants were identified using the Variant Effect Predictor (VEP) consequence ‘synonymous_variant’.

#### Selection of gene features

We partitioned genes into six mutually-exclusive annotations based on LOEUF scores (Karczewski *et al*., 2020) and cumulative mutation rate (i.e. the sum of gene-wise mutation rates across synonymous, Mis0, Mis1, Mis2, and PTV). Specifically, these annotations were: LOEUF quintile 1 and mutation rate quintile 1 (LOEUF1_mu1), LOEUF quintile 1 and mutation rate quintiles 2-5 (LOEUF1_mu2), LOEUF quintile 2, LOEUF quintile 3, LOEUF quintile 4, and LOEUF quintile 5. We stratified our analyses by LOEUF due to the well-understood enrichment of autism *de novo* association signal in constrained genes (Kosmicki *et al*., 2017). We stratified our analyses by mutation rate to avoid bias in model estimation due to mutation-rate-dependent architecture. We only stratified LOEUF quintile 1 by mutation rate because other LOEUF quintiles had very little association signal (**Figure 1**). We stratified mutation rate into quintile 1 and quintile 2-5 to roughly balance the number of observed mutations to preserve power.

#### Prevalence

We incorporated prevalences that vary as a function of three proband variables:

- Cohort: For SPARK and ASC, we used the most recent CDC estimate for the prevalence of autism, 0.0276 (Maenner et al, 2023). For GeneDX, reflecting the fact that this cohort is comprised of probands that are, on average, more severely affected, and referred for clinical genetic testing, we used a lower prevalence of 0.01. This choice is supported by Seplyarskiy, Moldovan et al (2025), who analyzed cohorts including GeneDX, examined the distribution of gene-wise rate ratios, and found that they very rarely exceeded 100, suggesting that 1/100 is a plausible lower bound for NDD prevalence. For analyses incorporating all three cohorts, we computed an average weighted by the cohort sample sizes, yielding an overall autism prevalence of 0.022.
- Sex: Recent CDC estimates suggest that the prevalence of autism in boys and girls is 0.0430 and 0.0114, respectively. For analyses stratifying males and females across all cohorts, we multiplied the cohort weighted average, 0.022, by (0.043/0.0276) to estimate sex-specific prevalences. For analyses stratifying by cohort and sex, we multiplied cohort-specific estimates by the same scaling factors.
- DD/ID: Recent CDC estimates suggest that 37.9% of ASD probands have DD/ID. For analyses stratifying by DD/ID, we multiplied the cohort weighted average by 0.379 and (1-0.379) to estimate prevalences of autism with and without DD/ID. For analyses stratifying by cohort, we multiplied the cohort-specific estimates by the same scaling factors. For analyses stratifying by sex, we used sex-specific DD/ID prevalence in autism estimates from the same CDC report (0.421 for girls, 0.369 for boys) to compute similar scaling factors. We note that the prevalence of DD/ID within autism in our analyses is taken from a contemporary epidemiological study of autism, which will apply to varying extents in our cohorts due to the fact that these cohorts were collected at varying points in the preceding decades.

#### Standard error calculation

To compute standard error of model coefficients and all downstream estimands, we use a gene bootstrap with 100 iterations.

### Mutation rate misspecification

Misspecification of mutation rate could lead to biased estimation of the distribution of *γ*_*g*_. In particular, mutation rates vary across the genome due to factors such as replication timing, recombination, and chromatin modification (Seplyarskiy *et al*., 2023), which can lead to some genes having mutation rates that differ from expectation based only on trinucleotide context and methylation. These features affect sites regardless of functional consequence. One approach to address this issue is to leverage an external population reference dataset to estimate the degree of mutation rate misspecification for each gene for a set of neutral variants, and scale the mutation rates appropriately. Conveniently, burdenMLE itself can be used to do this in two steps.

In step 1, we use burdenMLE to estimate the distribution of multiplicative mutation rate misspecification across genes:

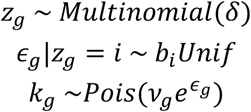

Where *k*_*g*_ is the observed number of polymorphic sites in the external dataset, *v*_*g*_ is the expected number of polymorphic sites, *N* is the number of individuals, and *ϵ*_*g*_ is the (log-scale) misspecification. In contrast to our main burdenMLE analyses, here we do not stratify by any gene-level features, and use mixture components with both positive and negative log rate-ratios (specifically, 31 mixture components ranging from -2 to 2). We fit this model to publicly available synonymous variant counts from the gnomAD v4 dataset (Chen *et al*., 2024).

In step 2, once the distribution of *ϵ*_*g*_ is inferred, the posterior mean is:

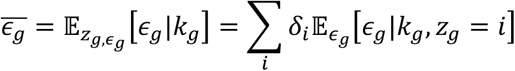

Where *δ*_*i*_ are the inferred mixture proportions. We approximate E_*ϵg*_]*ϵ*_*g*_|*k*_*g*_, *z*_*g*_ = *i*] with a discrete approximation to the *ϵ*_*g*_|*z*_*g*_ = *i*∼*b*_*i*_*Unif* distribution (see *Poisson-Uniform evaluation grid*) above. The modified burdenMLE observation model for *de novo* variants in autism is then:

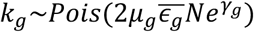

### Mutational variance

Traits and diseases can be modelled additively as the sum of genetic, environmental, and *de novo* mutational components

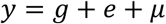

Variation in complex traits then is similarly composed of additive components:

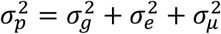

The mutational variance is the proportion of phenotypic variance that is attributable to *de novo* variants:

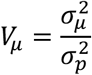

The mutational variance due to gene *g* is:

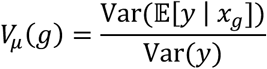

Where *x*_*g*_ is a random variable denoting the genotype. Because we analyze very rare mutations, we assume that *x*_*g*_ is binary, i.e. that each individual has 0 or 1 *de novo* mutations in gene *g*.

Expanding the term E[*y* | *x*_*g*_]:

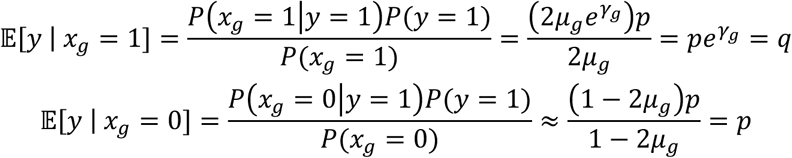

Where, in the second line, the approximation is accurate when most cases do not carry a mutation in gene *g*. The variance of E[*y* | *x*_*g*_] is then

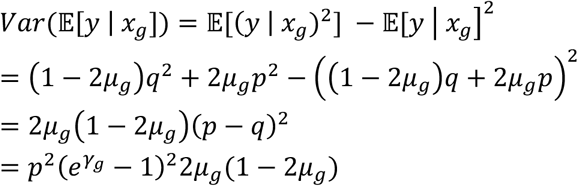

Which gives the following expression for *V*_*μ*_(*g*):

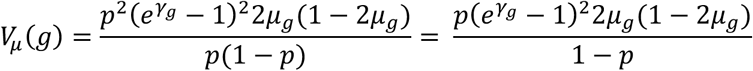

The mutational variance *V*_*μ*_ is then:

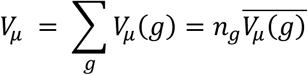

Where *n*_*g*_ is the number of genes and 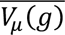 is the mean *V*_*μ*_(*g*) across genes. We considered two approaches to estimate *V*_*μ*_: (1) computing exact moments of the inferred effect size distribution, and (2) summing gene-wise posterior mean mutational variance. These two estimators will disagree when there is dependence between effect sizes (*γ*_*g*_) and mutation rates (*μ*_*g*_) that is not modelled by the gene annotations. We note that in our analyses, these two methods agree (i.e. the sum of PTV and Mis2 in **Figure 2B**, i.e. the exact method, and the cumulative sum in **Figure 2C**, i.e. the gene-wise posterior method).

Throughout the manuscript, we estimate *V*_*μ*_ on the observed (i.e. binary) scale.

### Other features of the *de novo* effect size distributions

In addition to aggregate mutational variance across genes, we also estimate gene-wise mutational variances, the number of genes with some minimum effect size *r*, the fraction of cases carrying a variant with some minimum effect size, and the effective penetrance of autism-associated *de novo* variants. For these quantities, we compute posterior means of the function of interest *f*(*γ*_*g*_) over uncertainty in both *z*_*g*_ and *γ*_*g*_:

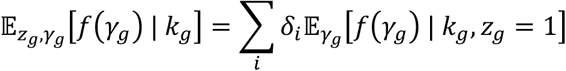

Where *δ*_*i*_ are the inferred mixture proportions. We approximate E_*γg*_]*γ*_*g*_ | *k*_*g*_, *z*_*g*_ = *i*] with a discrete approximation to the *γ*_*g*_|*z*_*g*_ = *i*∼*b*_*i*_*Unif* distribution (see *Poisson-Uniform evaluation grid*) above.

#### Gene-wise mutational variance

To compute gene-wise mutational variance, we compute the posterior mean of:

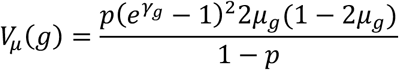

#### Number of genes with minimum effect size *r*

The number of genes with minimum effect size *r* is:

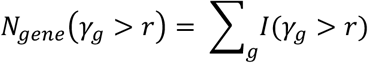

Where *I*(*γ*_*g*_ > *r*) is an indicator random variable encoding whether *γ*_*g*_ is greater than *r*. We compute the posterior means of *I*(*γ*_*g*_ > *r*) per gene and sum across genes to estimate *N*_*gene*_(*γ*_*g*_ > *r*). We used a similar approach to estimate the number of genes from previous gene-discovery efforts with minimum effect size *r* by summing *I*(*γ*_*g*_ > *r*) across significant genes (as in **Figure 3A**, bottom two rows), as well as the mean CDS of genes with rate ratio greater than *r* by weighting the mean CDS length across genes by these posterior means.

#### Fraction of cases carrying a variant with effect size greater than *r*

In clinical genetics, a quantity of interest is the proportion of cases who have a highly penetrant causal variant; such variants may be returned as genetic diagnoses.

For a gene *g* with effect size *γ*_*g*_, the fraction of cases carrying a variant in that gene:

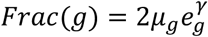

The fraction of cases with a variant with minimum effect size *r* (high penetrance, “HP”) is then:

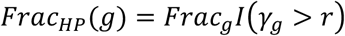

Where *I*(*γ*_*g*_ > *r*) is an indicator random variable encoding whether *γ*_*g*_ is greater than *r*. We compute the gene-wise posterior of this function for all genes. The aggregate fraction of cases carrying a variant with effect size greater than *r* is then:

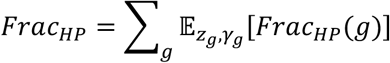

#### Effective penetrance

What is the average penetrance of an autism-causing *de novo* variant? The penetrance of mutations in gene *g* is:

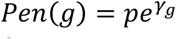

Which is interpretable as the frequency of *de novo* variants in cases. A naïve approach to estimating average penetrance would be to simply average penetrance estimates across all genes; however, this approach ignores the fact that the majority of genes are not expected to be causal for autism. To address this issue, we define the *effective penetrance* as the average penetrance weighted by excess in cases versus controls:

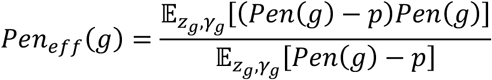

This quantity is interpretable as the average gene-wise penetrance of an excess *de novo* variant in autism cases versus controls. We compute the gene-wise posterior means of the numerator and denominator of this function across genes and average the resulting quantities weighted by mutation rate, to estimate the genome-wide effective penetrance:

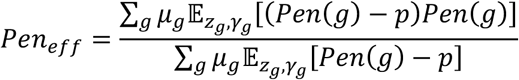

### Interpretation of burden effects

Throughout this work, we estimate and analyze gene-wise burden effect sizes, i.e. pooling variants within functional categories. For burden masks where variants have identical effect sizes (as is approximately true for PTVs), the burden and total (i.e. modelling individual variant-wise effects) genetic architecture are the same. For burden masks where variants have variable effect sizes (as is likely the case for missense variants), genetic architecture estimates from burden effects may differ from similar estimates using a model for variant-wise effects. We mitigate this attenuation by stratifying missense variants into three bins (see *burdenMLE implementation details* above), but such variability may remain within these bins.

Consider the scenario where, within a burden mask, half of effects are null and half have identical non-zero effects. In this setting, the burden mutational variance is equal to one half of the total mutational variance (Weiner, Nadig, et al, 2023). The estimated rate ratio will be less than the rate ratio estimated only from non-null alleles, which may change whether the gene is classified as having a “large effect”, and the fraction of cases estimated to be carrying “large effect” variants. In such a setting, the true effective penetrance is equal to the prevalence times the non-null effect size, but the burden effective penetrance will be equal to half this quantity.

### Short insertions and deletions

In addition to single nucleotide variants, short coding insertions and deletions (indels) are captured by exome sequencing and can cause gene loss-of-function. Modelling indels with burdenMLE is challenging, as mutation rates for indels have not been resolved as confidently as for single-nucleotide variants. We instead approximate the contribution of frameshift indels by scaling our loss-of-function SNV burdenMLE estimates by the ratio of loss-of-function variants (inclusive of frameshift indels and SNVs) to non-indel loss-of-function variants (i.e. SNVs only), which was 1.91 for the autism dataset and 1.98 for the DDD dataset. The quantities that we scaled with this factor are the loss-of-function estimates of mutational variance and fraction of cases with large-effect variants. We did not do the same scaling procedure for in-frame missense indels; these make up <5% of missense variants (Kwon *et al*., 2024), and they are not expected to be functionally equivalent to missense SNPs in the same gene. This choice is expected to produce a small amount of downward bias in our estimates of missense variant mutational variance.

### Dataset specific scaling parameters

A large trio sequencing study may be composed of several studies with differing ascertainment strategies. Different ascertainment strategies may lead to heterogeneity in estimated effect size, as rate ratios scale with the prevalence of the phenotype. To study this phenomenon, we estimated study- and sex-specific scaling parameters in three steps.

First, as described above, we analyze the aggregate dataset with burdenMLE, without stratifying by study and sex, to estimate the distribution of total dataset effect sizes *γ*_*g*_.

Second, we modelled dataset- and sex-specific effect sizes for study *j*, *γ*_*g,j*_, as being constant multiples of the total dataset effect size:

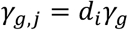

We developed a simple method to estimate these scaling factors that characterize whether a component dataset of our sample has a smaller or larger excess of *de novo* mutations than the total sample. The primary purpose of this approach is to increase power, which it does by leveraging the distribution inferred in the larger overall sample, and using the component datasets only to estimate the scaling factor. We applied this method to estimate study- and sex-specific scaling factors. This approach is entirely driven by the component dataset and total dataset mutation counts, does not explicitly incorporate information about how the component datasets are defined, and is not subject to bias by component dataset sample size. This approach is designed to capture differing ascertainment severity, and is appropriate when the scale, and not the pattern of effects across genes, varies across component datasets, which is true when component datasets vary only in prevalence of the underlying phenotype (e.g., the same phenotype with different average severity).

Specifically, given that a particular gene has *k*_*g*_ *de novo* variants in the total dataset, the number of *de novo* variants in a particular dataset follows a binomial distribution:

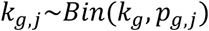

Where *p*_*j*_ is the ratio of expected mutation counts between dataset *i* and the total dataset:

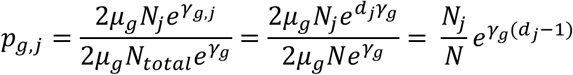

Where *N*_*j*_ and *N* are the dataset-specific and total sample sizes, respectively.

The maximum-likelihood estimate of *d*_*i*_ is

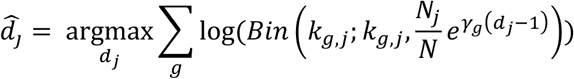

To evaluate the log likelihood, we integrate this function over the posterior distribution of *γ*_*g*_ as described above. We compute *d̂*_*y*_ using a grid search over plausible values of *d*_*j*_ (specifically, ranging from 0.65 to 1.35). Standard errors were computed using the observed Fisher information, estimated numerically as the second derivative of the log-likelihood function via a central difference approximation.

Lastly, we used these maximum likelihood estimates of *d*_*j*_ to estimate study- and sex-specific mutational variances and cases carrying a variant with effect size greater than *r* (**Figure 2G,H**). We also used these estimated scaling factors to forecast variability of gene discovery across ascertainment strategies (see *Forecasting gene discovery*).

### Forecasting gene discovery

To estimate gene discovery at varying *N*, we simulated data from our inferred burdenMLE models at varying sample size and computed the number of significant discoveries. In detail, for each gene, we sampled an effect size from its fitted gene-wise posterior distribution, conditional on its observed de novo count, mutation rate, and annotations, and retained the gene’s observed mutation rate when simulating future data. Using these simulated effect sizes and mutation rates, we sampled counts *k*_*g*_ for each gene from the Poisson observation model *Pois*(2*μ*_*g*_*Ne*^*d*(*γg*^) for a particular choice of sample size *N* and study-specific scaling factor *d*_*j*_. We computed p-values by evaluating the cumulative distribution function of the null mutational distribution *Pois*(2*μ*_*g*_*N*) at *k*_*g*_ (i.e. against the null hypothesis that *γ*_*g*_ = 0).

For each simulation, we report the number of significant genes after Benjamini-Hochberg (Benjamini and Hochberg, 1995) and Bonferroni correction. We ran 100 simulations for 20 values of *N* ranging from 39680 (i.e. 1000 more samples than the current sample size of 38680) to 77360 (i.e. twice the current sample size of 38680). We note that this inference procedure is different, and in particular much simpler than, the TADA method that is commonly used for autism gene discovery; we choose this simpler test because the TADA method integrates several classes of evidence including *de novo* CNVs and case/control comparisons, making it difficult to analyze with burdenMLE. For these analyses, we used the inferred distribution of PTV effect sizes.

### Simulations

We conducted simulations to evaluate the performance of burdenMLE across different distributions of *γ*_*g*_ and sample sizes. Using empirical PTV per-gene mutation rates, we simulated case counts under four distinct effect size distributions:

*Mixture of half-uniforms*: The burdenMLE generative model. A mixture model with 10 components, where effect sizes were drawn from half-uniform distributions with varying endpoints (0 to log(100)), with mixture weights drawn from a Dirichlet distribution.
*Half-Infinitesimal:* A positive half-normal distribution with standard deviation uniformly sampled between 0.5 and 1.0 for each simulation.
*Point-Normal*: A mixture distribution where a fraction (sampled uniformly between 0.85 and 0.95) of genes had effect size of 0, while the remaining genes had effect sizes drawn from a normal distribution with mean sampled uniformly between 1.5 and 2.5 and standard deviation sampled uniformly between 0.2 and 0.4.
*Oligogenic*: A distribution where a small proportion of genes (sampled uniformly between 0.1% and 1%) had large effects (sampled uniformly between log(5) and log(50)), while all other genes had no effect.

For the Half-Infinitesimal, Point-Normal, and Oligogenic simulations, the parameter choices led to a range of mutational variances, mostly between 0 and 0.04. For the mixture of half-uniform simulation, the parameter choices led to mutational variances ranging from 0 to 1; we did so to explore estimation accuracy in the full range of parameter values under the burdenMLE generative model.

For each architecture, we simulated data across five sample sizes (1,000, 5,000, 15,000, 38,680, and 50,000 trios) with 100 replicates per condition. Case counts were generated using the Poisson observation model *Pois*(2*μ*_*g*_*Ne*^*γg*^). For each simulation, we used burdenMLE to estimate the total mutational variance, the fraction of cases carrying a variant with effect size greater than 5, and effective penetrance.

### Autism exome sequencing data

We used counts from the latest freeze of trio exome sequencing data from the Autism Sequencing Consortium (ASC) (N = 38680). Briefly, this dataset is composed of large samples drawn from four sources: (1) The Simons Powering Autism Research (SPARK) study, a large national study (SPARK Consortium, 2018) that recruited individuals who self-reported autism diagnoses, (2) the Autism Sequencing Consortium, a combination of datasets from multiple studies (Buxbaum *et al*., 2012), which vary in ascertainment but generally used expert-rated diagnostic instruments, (3) samples from GeneDX, a clinical genetics testing company, where individuals were referred by a physician for clinical genetic testing, and (4) individuals with autism from the Deciphering Developmental Disorders cohort (Kaplanis *et al*., 2020). Data collection, alignment, and variant calling are described in Satterstrom et al (2026)

Briefly, for datasets previously analyzed in Fu et al., autosomal calls were taken directly from published supplementary data, while sex chromosome variants were re-called using a modified Hail-based de novo caller capable of handling hemizygous regions. For additional datasets (ASC B17–21, SPARK iWES v2, and GeneDx), de novo variants were identified using a comparable pipeline, including genotype quality filtering and likelihood-based calling, with dataset-specific adaptations where necessary.

Across all datasets, raw calls were subjected to stringent quality control filters, including rarity thresholds based on gnomAD and within-dataset allele frequency, allele balance and depth constraints, and strand bias metrics. Additional dataset-specific filters were applied to ensure high-confidence calls, reflecting differences in sequencing platforms and cohort size. Following filtering, variants were functionally annotated, and analyses were restricted to at most one de novo variant per gene per individual, prioritizing the most severe predicted consequence. Samples with an excess burden of coding de novo variants relative to expectation were excluded. A detailed description of the de novo calling pipeline can be found in most recent flagship ASC manuscript (Satterstrom et al, 2026).

Recorded DD/ID status was defined differently across cohorts, as described in Satterstrom et al (2026). ASC probands were recorded with DD/ID when the contributing site reported intellectual disability or IQ <70, the prevalence of which was 23%. SPARK probands were recorded with DD/ID based on a reported professional diagnosis of intellectual disability, cognitive impairment, global developmental delay, or borderline intellectual functioning, the prevalence of which was 16%. GeneDx probands were recorded with DD/ID based on HPO terms, the prevalence of which was 71%. These different definitions could exaggerate the difference in DD/ID prevalence between cohorts.

### Developmental disorder exome sequencing data

We used published *de novo* mutation counts from Kaplanis et al (Kaplanis *et al*., 2020), which sampled 31058 trios from three sources: (1) the Deciphering Developmental Disorders project, a large collection from the UK (Firth, Wright and DDD Study, 2011), (2) Radboud University Medical Center, and (3) GeneDx, a clinical genetics company. We annotated variants from this dataset in an identical manner to the autism sample (i.e. into PTV, Mis2, Mis1, Mis0, and synonymous). We note that 3543 samples with co-morbid developmental disorder and autism in the GeneDx dataset are shared between our analyses of developmental disorder and autism.

### Paternal Age Analysis

We evaluated whether differences in paternal age could meaningfully influence the estimated mutational variance. Paternal age at the proband’s birth was available for 17,871 of 18,060 SPARK probands (99.0%) and was compared with paternal ages reported in the 2024 CDC/NCHS natality records. We estimated the relationship between paternal age and de novo mutation rate within SPARK using a Poisson regression of each proband’s synonymous de novo variant count on paternal age, adjusting for sequencing dataset and proband sex. The fitted model estimated a 2.26% increase in mutation rate per additional paternal year (95% CI, 1.83%–2.68%). Mean paternal age was 32.54 years in SPARK and 32.13 years among U.S. births with reported paternal age. Combining these estimates of paternal age difference and rate of mutation accumulation, we estimate that our mutation rate estimates may be 0.92% higher in SPARK (95% CI, 0.75%–1.10%). Because mutational variance scales linearly with mutation rate, we assessed sensitivity by dividing the estimated PTV and damaging-missense mutational variances by this mutation-rate ratio. This adjustment reduced the combined estimate from 0.0211 to 0.0209, indicating a negligible expected effect of paternal age

## Code availability

Code for running burdenMLE-DN is available at https://github.com/ajaynadig/burdenMLE-DN, including a wiki and tutorial.

