## Supplementary Figures for "Estimating the contribution of coding mutations to autism"

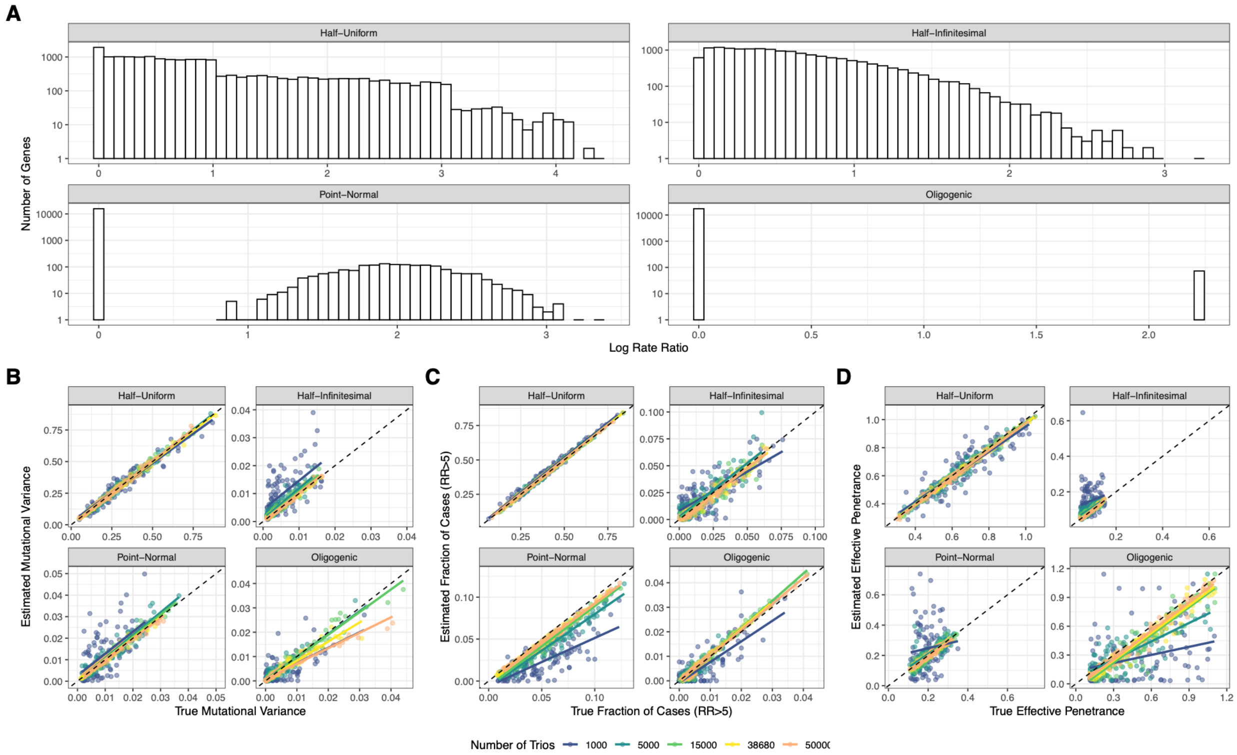

**Supplementary Figure 1. Full simulation results.** We simulated four effect-size distribution families at five sample sizes, with 100 replicates per combination and simulation-specific hyperparameters. **(A)** Example distributions of gene-level log rate ratios under the half-uniform, half-infinitesimal, point-normal, and oligogenic architectures. **(B)** Comparison of true and estimated mutational variance. **(C)** Comparison of the true and estimated fraction of cases carrying a large-effect mutation (rate ratio > 5). **(D)** Comparison of true and estimated effective penetrance. Colors denote sample size, and dashed lines denote equality between the true and estimated quantities. Numerical results are provided in Supplementary Table 1.

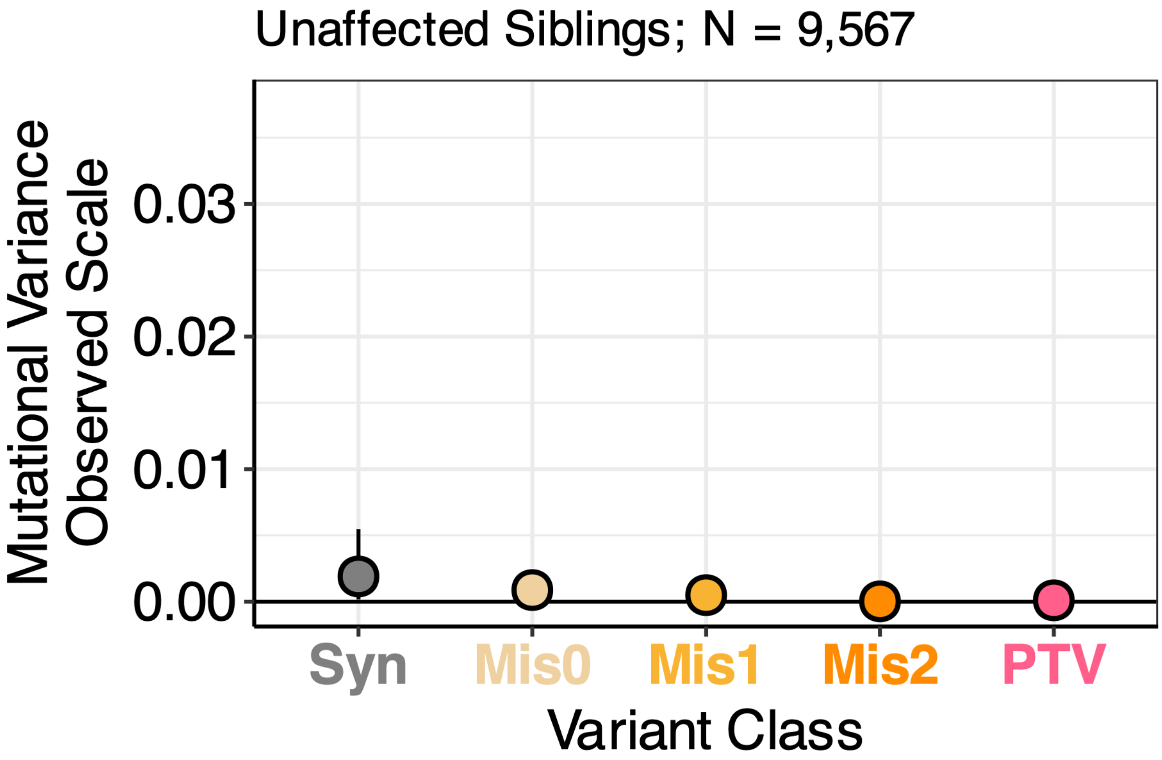


**Supplementary Figure 2. Mutational variance estimates in unaffected siblings.** Estimates are shown for 9,567 unaffected siblings using an assumed autism prevalence of 2.22%. Points indicate estimates and error bars indicate gene-bootstrap 95% confidence intervals.

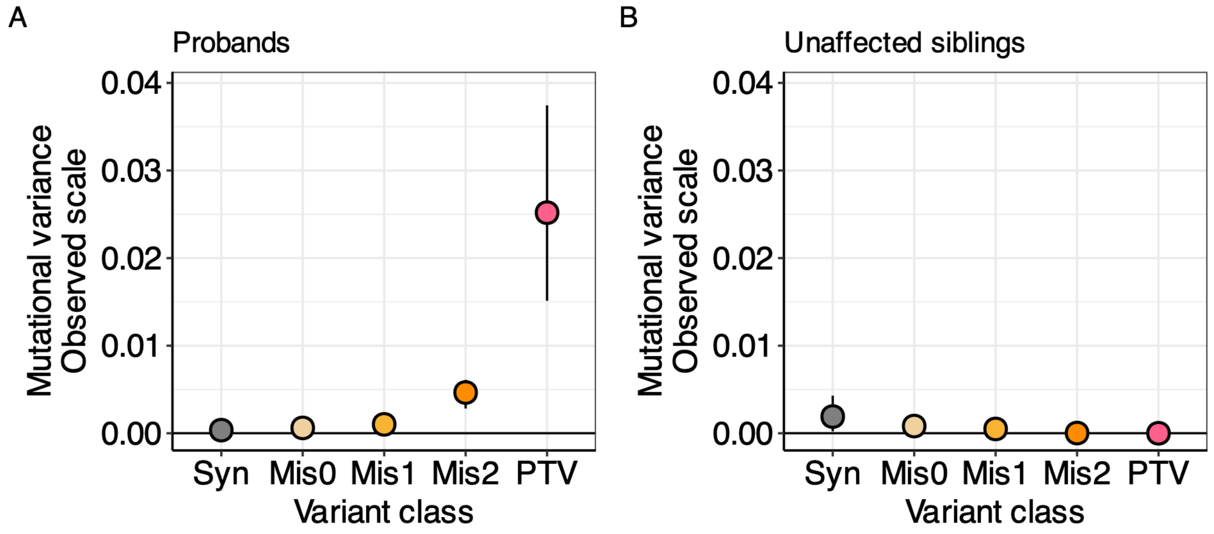


**Supplementary Figure 3. Exclusion of genes with evidence of clonal expansion in spermatogonia.** Clonal expansion in spermatogonia can produce an excess of observed de novo mutations relative to mutation-rate expectations and is therefore a potential confounder in trio studies. We excluded the 69 clonal-expansion genes identified by Seplyarskiy et al. and re-estimated mutational variance in **(A)** autism probands and **(B)** unaffected siblings. The same y-axis scale is used in both panels. Points indicate estimates and error bars indicate gene-bootstrap 95% confidence intervals.

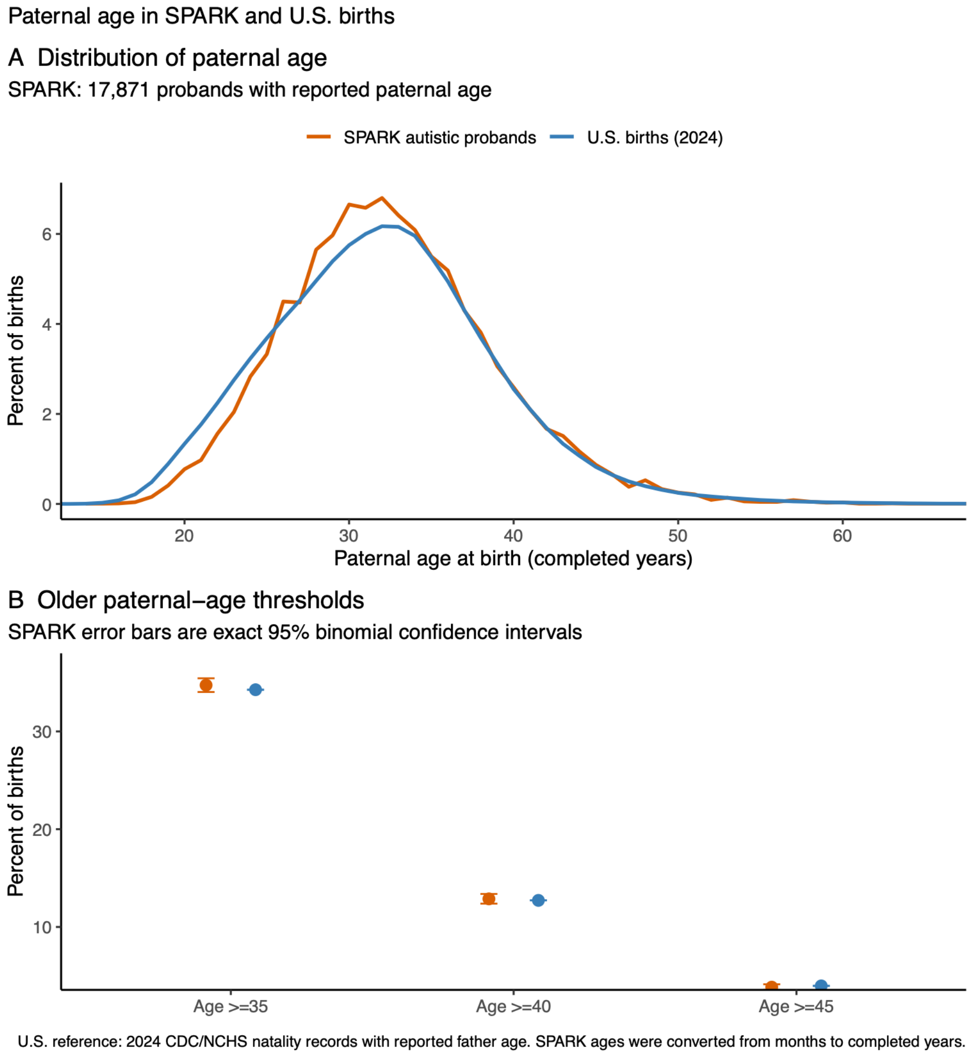


**Supplementary Figure 4. Paternal age in SPARK and U.S. births. (A)** Distribution of paternal age among 17,871 SPARK autistic probands with available paternal age and among U.S. births with reported paternal age in the 2024 CDC/NCHS natality records. SPARK ages, originally recorded in months, were converted to completed years. **(B)** Percentages of births with paternal age at least 35, 40, or 45 years. Error bars for SPARK indicate exact 95% binomial confidence intervals; the CDC/NCHS values represent the corresponding population proportions.

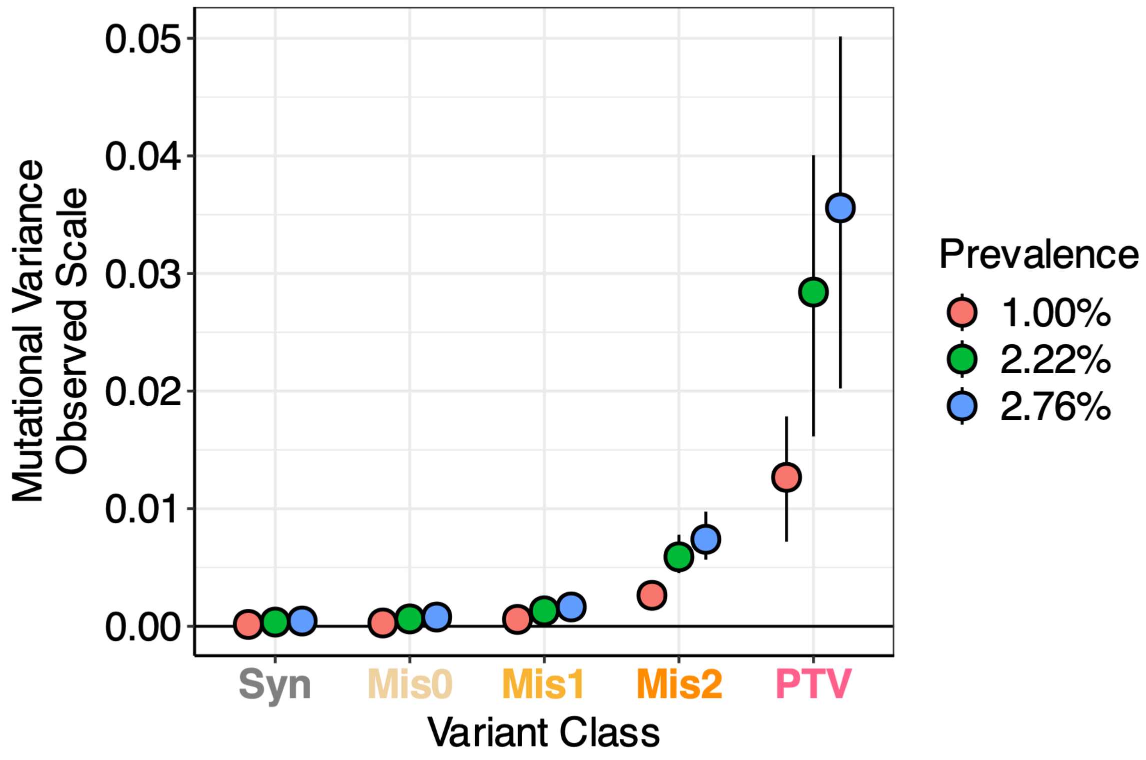


**Supplementary Figure 5. Mutational variance estimates across assumed autism prevalences.** We evaluated prevalences of 1.00%, 2.22%, and 2.76%. A prevalence of 2.76% approximates the prevalence of autism in the United States, 1.00% approximates the prevalence of autism with comorbid intellectual disability, and 2.22% is the sample-size-weighted value used in the primary combined-cohort analysis. The assumed prevalence p does not affect the estimated distribution of rate ratios, but mutational variance is proportional to p/(1-p). Points indicate estimates and error bars indicate gene-bootstrap 95% confidence intervals.

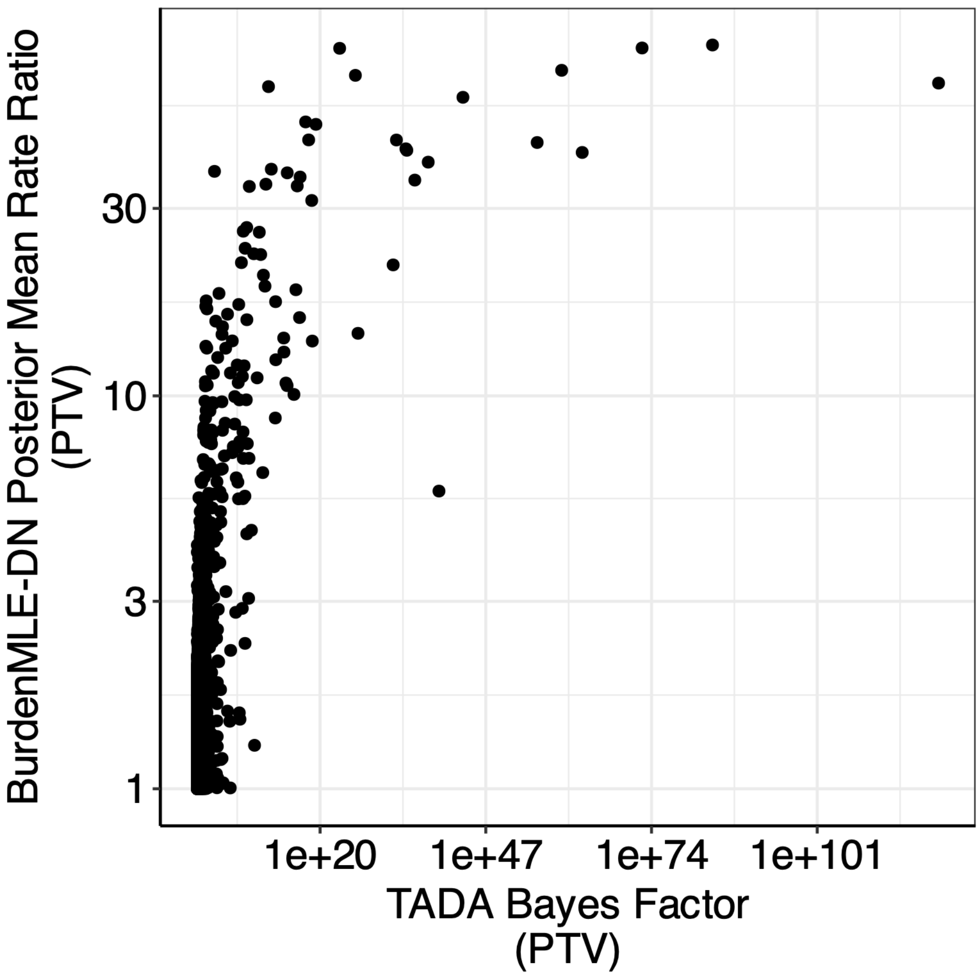


**Supplementary Figure 6. Comparison of BurdenMLE-DN and TADA evidence for autism-associated genes.** For each gene, the posterior-mean PTV rate ratio from BurdenMLE-DN is plotted against the PTV Bayes factor from the most recent Autism Sequencing Consortium TADA analysis. Both axes are shown on logarithmic scales.

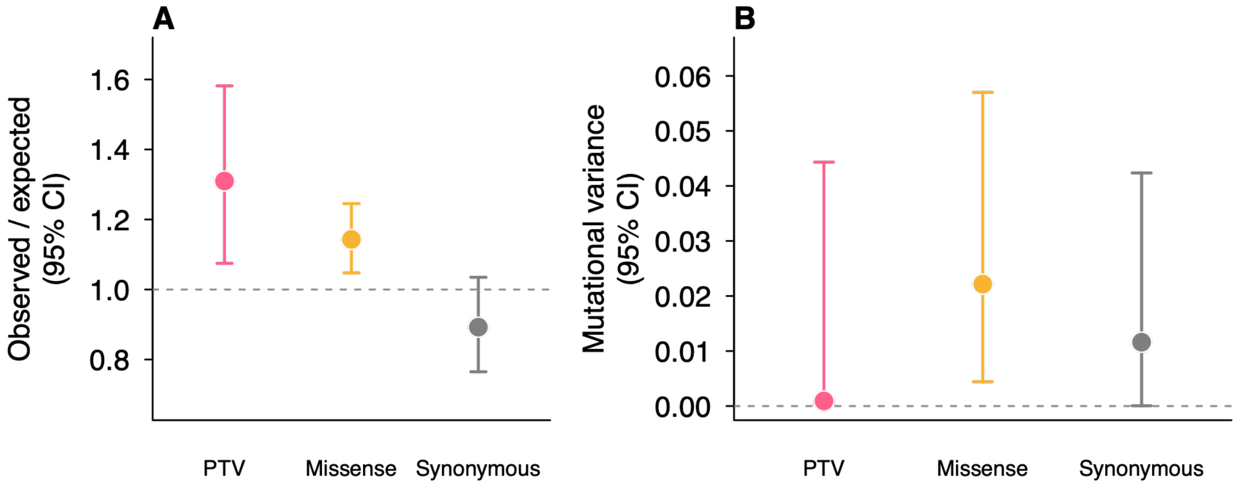


**Supplementary Figure 7. Application of BurdenMLE-DN to a Korean autism whole-genome sequencing dataset.** The analysis included 680 affected offspring. **(A)** Observed-to-expected mutation-count ratios for PTV, missense, and synonymous variants; error bars indicate exact 95% Poisson confidence intervals. **(B)** Observed-scale mutational variance estimates using an autism prevalence of 2.76%; error bars indicate gene-bootstrap 95% confidence intervals. PTV quantities include both SNVs and indels using the study-specific PTV indel scaling factor.

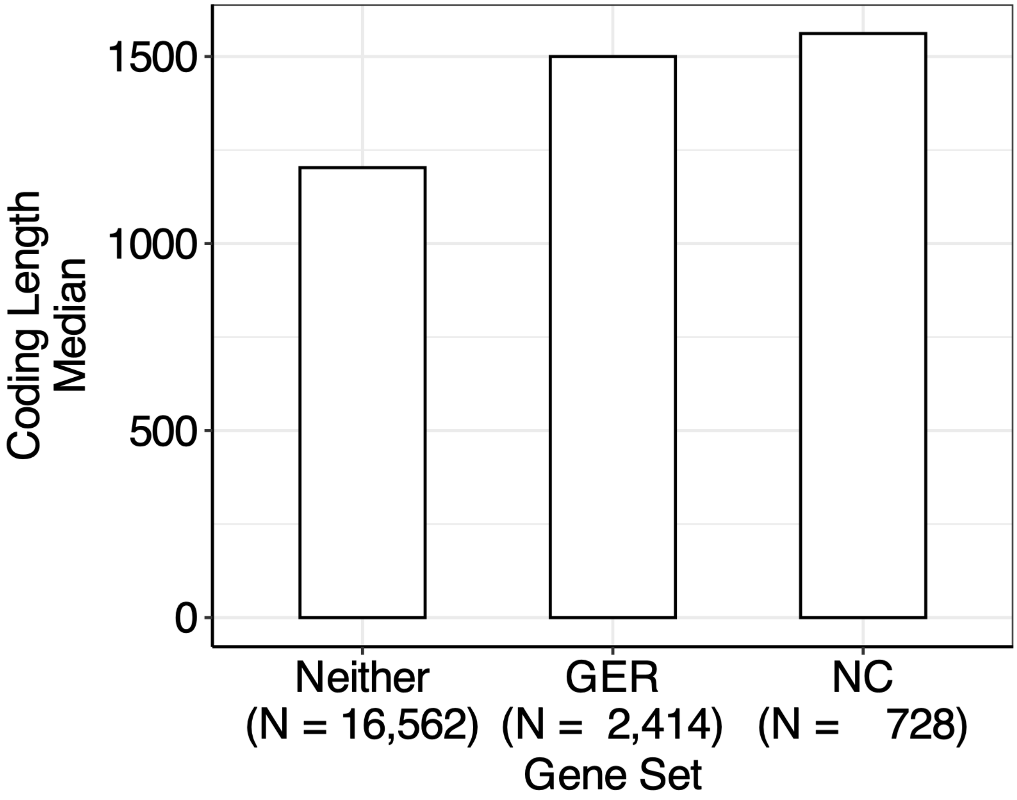


**Supplementary Figure 8. Coding sequence lengths of autism-relevant gene sets.** Median coding sequence length is shown for genes involved in gene-expression regulation (GER; GO:0006357), neuronal communication (NC; GO:0007268), or neither gene set. Gene sets were obtained from the AmiGO 2 database. Sample sizes indicate the number of genes in each group.

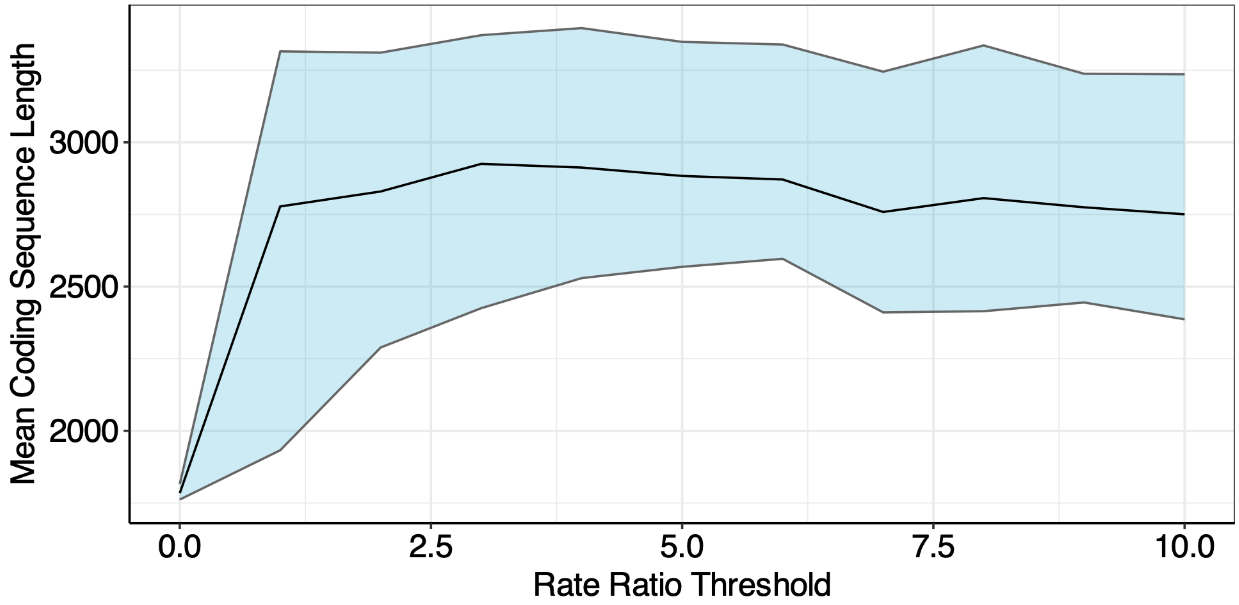


**Supplementary Figure 9. Coding sequence length across PTV rate-ratio thresholds.** For each threshold, the mean coding sequence length was calculated by weighting each gene by its posterior probability of having a PTV rate ratio greater than the threshold. The line indicates the estimate and the shaded region indicates the gene-bootstrap 95% confidence interval.

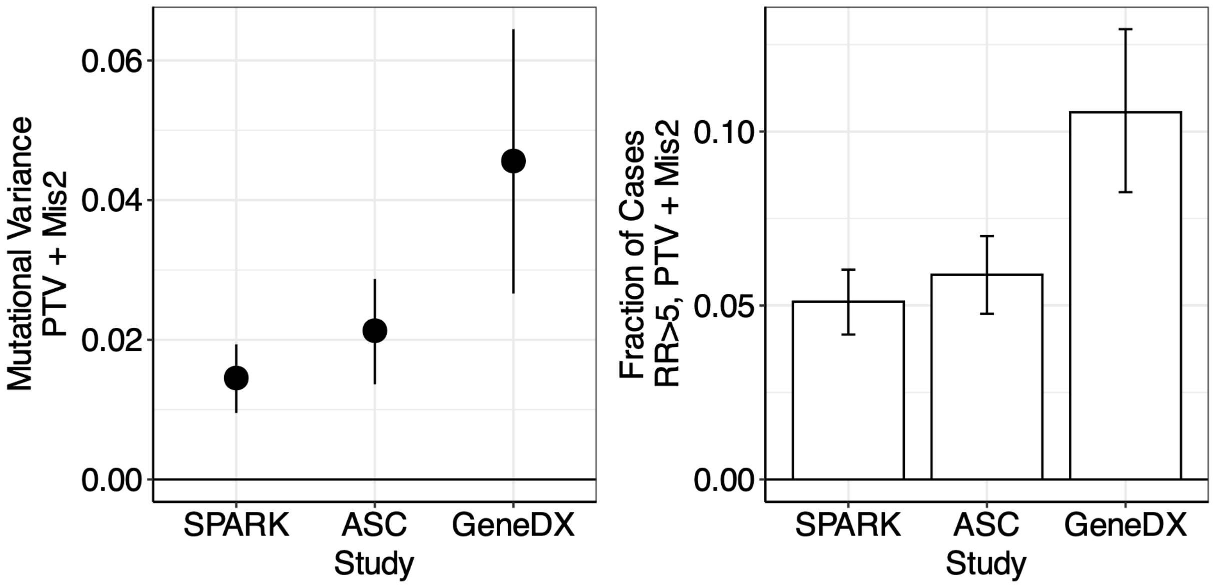


**Supplementary Figure 10. Heterogeneity in autism genetic architecture across cohorts.** Combined PTV and Mis2 mutational variance (left) and the fraction of cases carrying a PTV or Mis2 variant with rate ratio > 5 (right) are shown for SPARK, ASC, and GeneDx. Estimates use cohort-specific prevalence values. Points or bars indicate estimates and error bars indicate gene-bootstrap 95% confidence intervals.

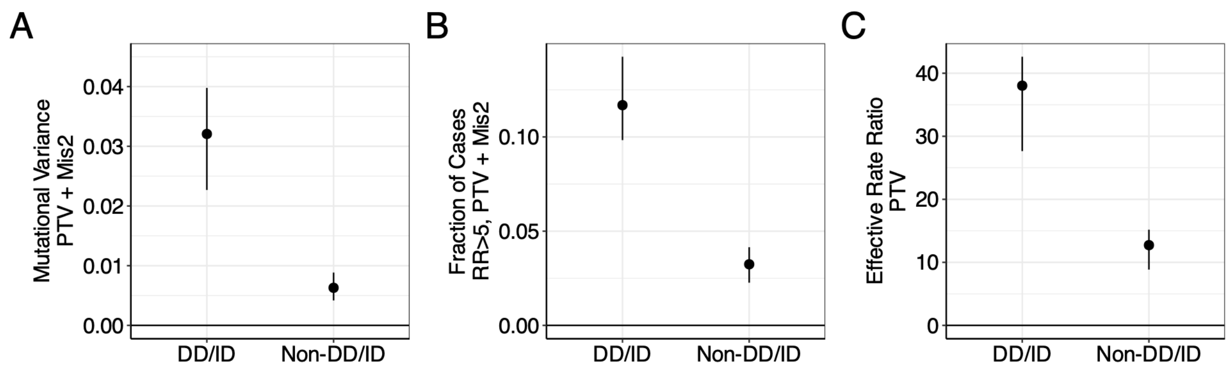


**Supplementary Figure 11. Autism genetic architecture according to comorbid developmental delay or intellectual disability.** Combined-cohort estimates are compared between autistic probands with DD/ID and those without DD/ID. **(A)** Combined PTV and Mis2 mutational variance. **(B)** Fraction of cases carrying a PTV or Mis2 variant with rate ratio > 5. **(C)** Effective PTV rate ratio. Points indicate estimates and error bars indicate gene-bootstrap 95% confidence intervals.

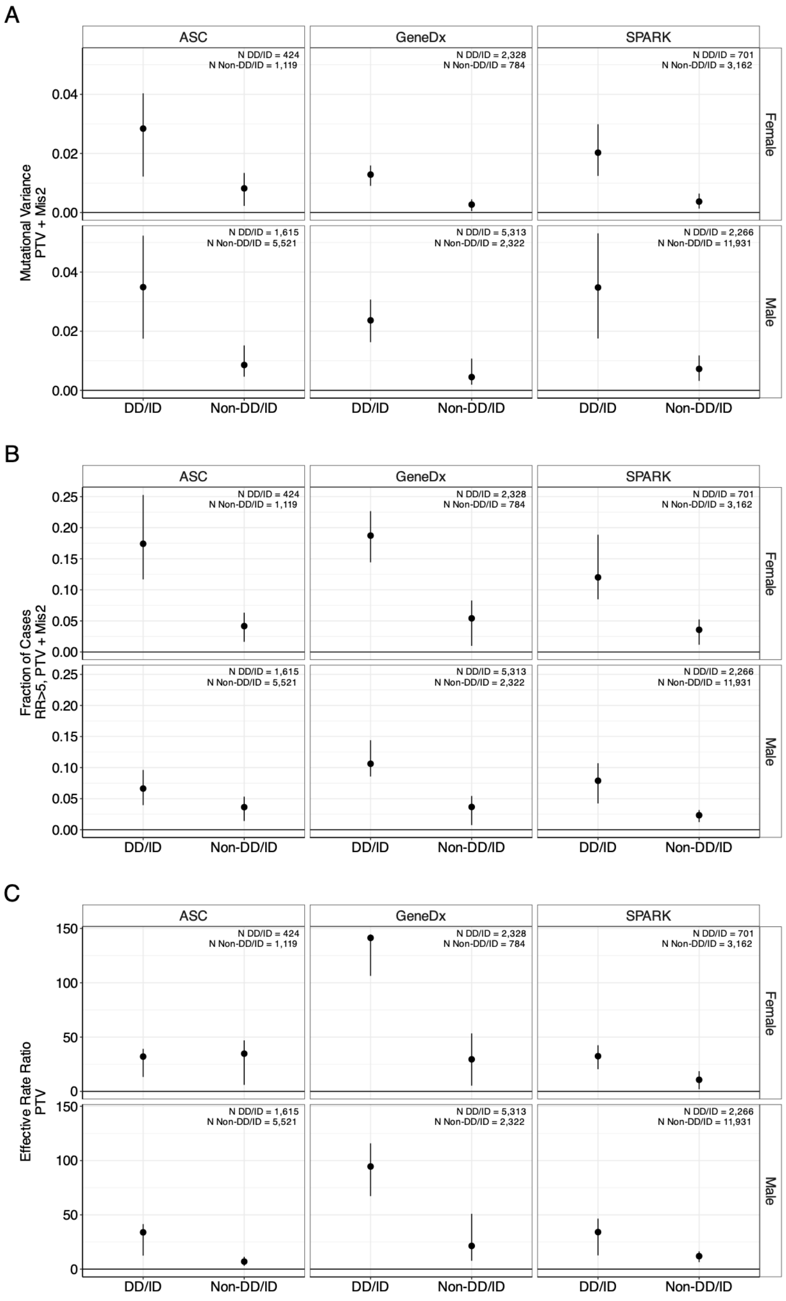


**Supplementary Figure 12. Autism genetic architecture jointly stratified by cohort, sex, and DD/ID.** Estimates are shown for ASC, GeneDx, and SPARK, separately for female and male probands with or without comorbid developmental delay or intellectual disability. **(A)** Combined PTV and Mis2 mutational variance. **(B)** Fraction of cases carrying a PTV or Mis2 variant with rate ratio > 5. **(C)** Effective PTV rate ratio. Points indicate estimates and error bars indicate gene-bootstrap 95% confidence intervals. The fine-grained strata have limited sample sizes and correspondingly imprecise estimates.

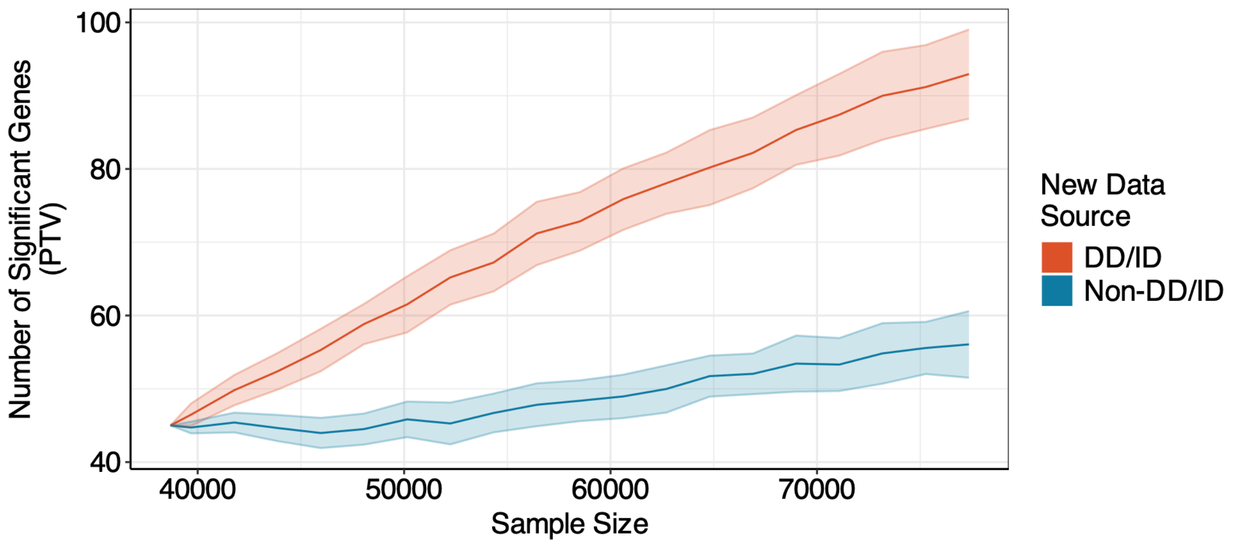


**Supplementary Figure 13. Forecasted autism gene discovery according to DD/ID ascertainment.** The number of genes expected to reach Bonferroni significance through de novo PTV association is shown as the total sample size increases, assuming that newly added probands either have DD/ID or do not have DD/ID. Lines indicate the mean across 100 simulations, and shaded regions indicate one standard deviation above and below the mean.

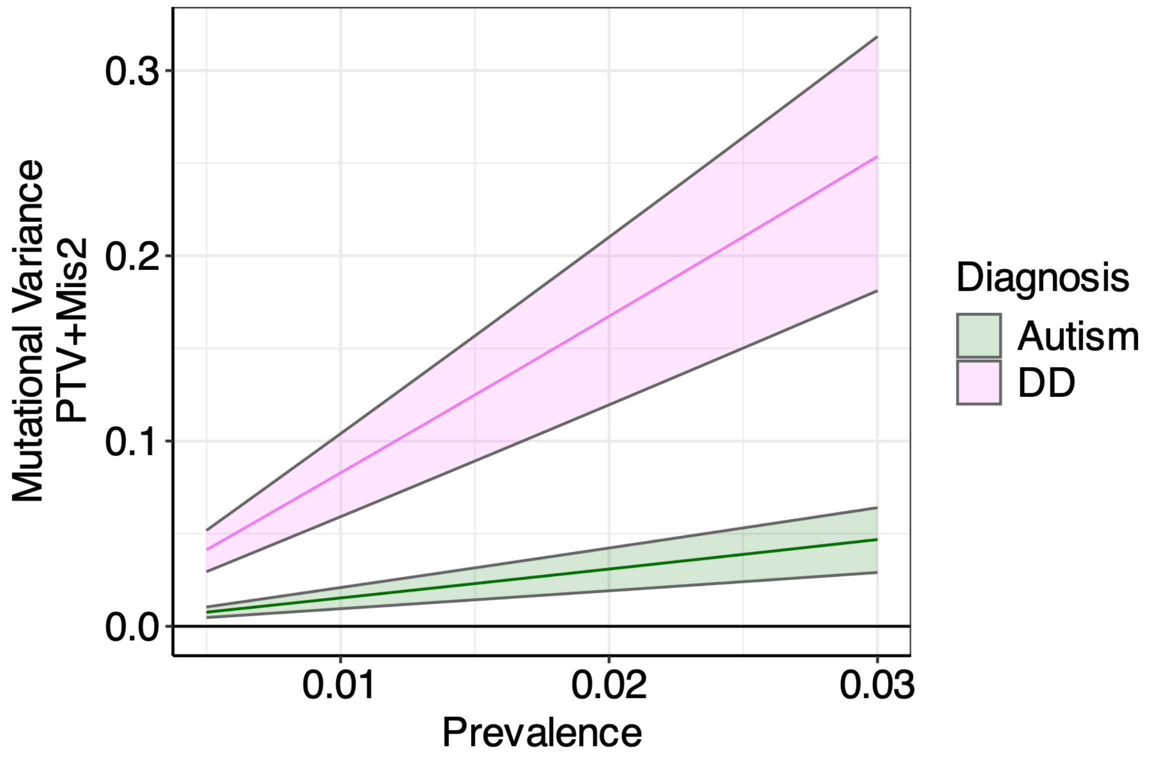


**Supplementary Figure 14. Sensitivity of autism and developmental-disorder mutational variance to assumed prevalence.** Combined PTV and Mis2 observed-scale mutational variance is shown across a range of assumed prevalences for autism and developmental disorders. Lines indicate estimates and shaded regions indicate gene-bootstrap 95% confidence intervals.

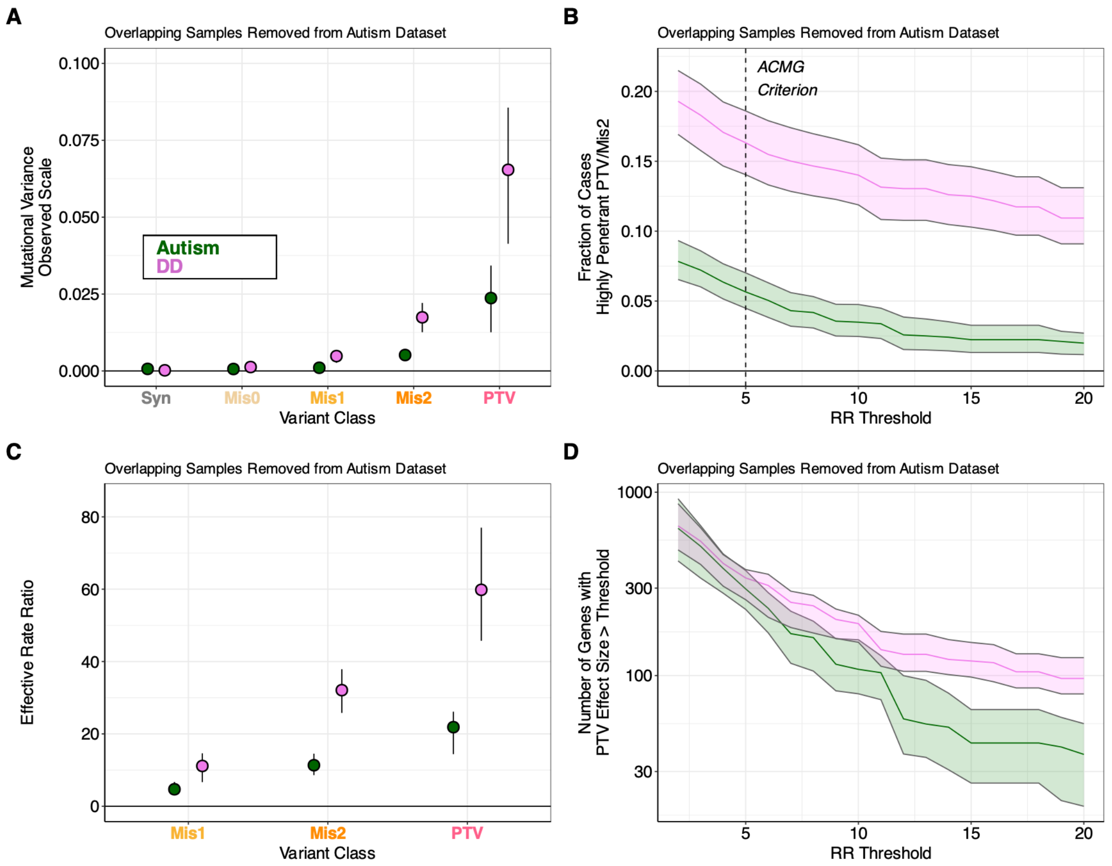


**Supplementary Figure 15. Autism and developmental-disorder estimates after removing overlapping samples from the autism dataset.** The 3,543 probands shared between the autism and developmental-disorder datasets were removed from the autism analysis and retained in the developmental-disorder analysis. **(A)** Observed-scale mutational variance by variant class. **(B)** Fraction of cases carrying a PTV or Mis2 variant above each rate-ratio threshold. **(C)** Effective rate ratio by variant class. **(D)** Number of genes with PTV rate ratio exceeding each threshold. Points and lines indicate estimates; error bars and shaded regions indicate gene-bootstrap 95% confidence intervals.

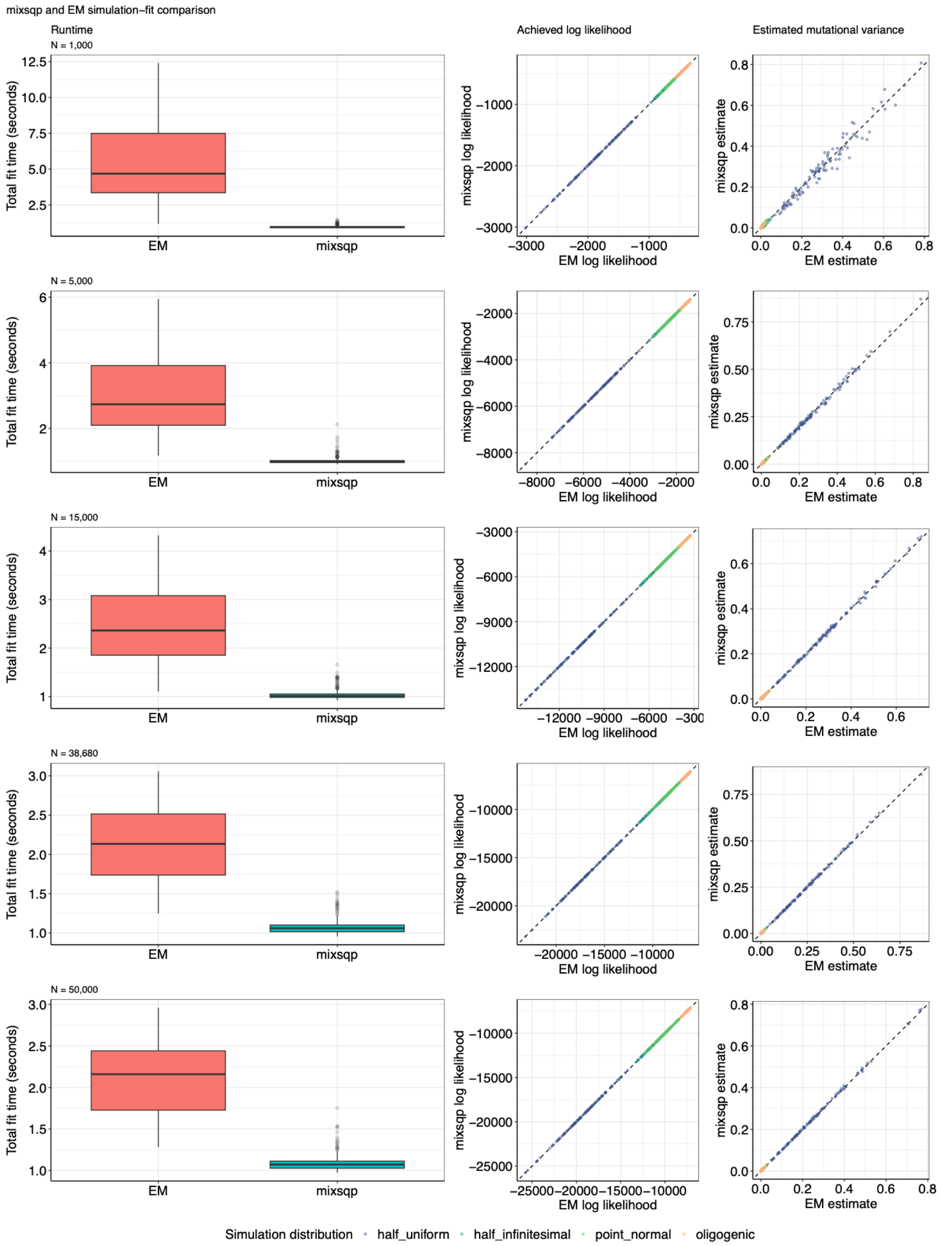


**Supplementary Figure 16. Comparison of mix-SQP and expectation-maximization optimization in simulations.** Rows correspond to simulated sample sizes of 1,000, 5,000, 15,000, 38,680, and 50,000 trios. The left column compares total model-fitting time, the middle column compares the achieved log likelihood, and the right column compares estimated mutational variance. Points in the paired comparisons are colored by simulated effect-size distribution; dashed lines denote equality between optimizers. Runtime comparisons summarize fits without bootstrap resampling.
